# Total plasma cfDNA methylation in kidney transplant recipients provides insight into acute allograft rejection pathophysiology

**DOI:** 10.1101/2024.08.17.24312147

**Authors:** Benjamin L. Spector, Boryana S. Koseva, Drinnan Sante, Warren A. Cheung, Reid S. Alisch, Alexander Kats, Phillip Bergmann, Elin Grundberg, Gerald J. Wyckoff, Laurel K. Willig

**Author notes:** **Corresponding author:** Benjamin L. Spector, MD, MS Clinical Sciences Center 600 Highland Ave Madison, WI 53792.

## Abstract

**Background:** Acute rejection threatens kidney allograft longevity. Cell-free DNA (cfDNA) is a real-time marker of organ injury and immune response. Donor-derived cfDNA (dd-cfDNA) has been leveraged as a biomarker of rejection, however, its reliability as a screening tool is unclear. DNA methylation is an epigenetic marker that informs regulatory element activity. We aim to elucidate differential methylation of total plasma cfDNA derived from pediatric kidney transplant recipients in the presence compared to absence of acute rejection. In doing so, we hope to exploit the property of cfDNA as a real-time biomarker and build on available testing to identify genes and processes participating in acute rejection pathophysiology in kidney transplantation.

**Methods:** Twenty plasma cfDNA samples from pediatric kidney transplant recipients were collected at the time of allograft biopsy. Using whole genome bisulfite sequencing, differentially methylated CpG residues (≥20% difference in methylation rate, *q*-value <0.05) were identified in presence (N = 7) vs absence (N = 9) of acute rejection. Separate analyses were performed comparing those with borderline rejection (N = 4) to those with rejection, and to those without rejection. Differentially methylated cytosines were then assessed for gene associations and pathway enrichments.

**Results:** In the comparison of acute rejection to non-rejection samples, there were 34,356 differentially methylated cytosines corresponding to 1,269 associated genes, and 533 enriched pathways. These numbers were all substantially greater (4-13x) than the comparisons made between acute rejection against those with borderline rejection, and between non-rejection against borderline rejection. Prominently enriched pathways between samples of individuals with and without acute rejection were related to immune cell regulation, inflammatory response, lipid metabolism, and tryptophan-kynurenine metabolism.

**Conclusions:** Our data suggest methylation plays a role in development of or response to acute kidney allograft rejection. Specifically, differentially methylated pathways associated with acute rejection include those related to immune and inflammatory responses.

## Introduction

End-stage kidney disease (ESKD) affects nearly 6,000 pediatric patients across the United States.^1^ The preferred treatment of ESKD is kidney transplant, however, allograft survival averages only 10-20 years.^2,3^ Factors such as exposures to nephrotoxic medications, chronic allograft nephropathy (CAN), and acute allograft rejection (AAR) threaten graft longevity. AAR increases risk for development of CAN, graft loss, and death.^4^ Enhanced understanding of molecular mechanisms of AAR has led to improved detection and monitoring of AAR through innovative assays including measurements of donor specific antibodies, urinary and plasma biomarkers, and donor-derived cell-free DNA (dd-cfDNA).^5,6^

Plasma dd-cfDNA assays are now clinically available as adjunct tools for diagnosing allograft injury. Under normal circumstances the majority of plasma cfDNA is derived from leukocytes.^7^ However, a relatively increased proportion of circulating cfDNA is generated by specific tissues and cells experiencing active injury.^7^ Dd-cfDNA assays attempt to leverage this relative increase in plasma dd-cfDNA to predict AAR, however, their utility as screening tools are unclear, with reported sensitivities for detecting AAR varying from 59-100%, depending on study and rejection subtype.^8–10^ Further, these assays struggle to differentiate AAR from other forms of acute kidney injury, and do not offer mechanistic insight into the molecular changes induced by AAR.^7^

DNA methylation is cell-specific and informs regulatory element activity and gene expression. As such, differential methylation of DNA loci allows identification of tissue of origin for a given DNA molecule, and provides insights into how disease states alter genetic regulation.^7^ In patients initiating kidney replacement therapy (KRT), whole blood samples collected at time of transplant or initiation of dialysis show differential methylation of genes involved in inflammation, vascular calcification, and cellular aging when compared to controls. However, at several months following initiation of KRT these DNA methylation changes are partially reversible.^11^ In kidney tissue, inflammatory genes and chronic kidney disease (CKD) markers are differentially methylated in the presence of allograft fibrosis.^12^ To date, the role of DNA methylation signatures in acute rejection of the kidney allograft is relatively underrepresented in the literature, however one study identified hypermethylation of genes enriched in immunologic pathways in peripheral blood mononuclear cells.^13^

In this study, we aim to elucidate differential DNA methylation levels of total plasma cfDNA in pediatric kidney transplant recipients in the presence as compared to absence of AAR. In doing so, we hope to exploit the property of cfDNA as a real-time biomarker and build on clinically available testing to identify potential genes and processes participating in rejection pathogenesis.

## Methods

### Regulatory Oversight

Prior to study initiation, approval was obtained from the Children’s Mercy Hospital Institutional Review Board under application STUDY00000188. Prior to study enrollment, informed consent was obtained from all participants aged 18-years and above; in the case of individuals younger than aged 18-years informed consent was provide by a parent/guardian. The research being reported is consistent with the Principles of the Declaration of Helsinki. The clinical and research activities being reported are consistent with the Principles of the Declaration of Istanbul as outlined in the “Declaration of Istanbul on Organ Trafficking and Transplant Tourism”.

#### Sample characteristics

Plasma was collected from pediatric kidney transplant patients at a single center (Children’s Mercy Hospital) at the time of clinically indicated kidney biopsy, which included for cause and surveillance biopsies. Surveillance biopsies were protocolized to take place at 6-, 12-, and 24-months post-transplant. Samples were then categorized according to their histopathologic diagnosis, as determined by a single pediatric nephropathologist (AK), as being non-rejection, borderline rejection, or acute allograft rejection based on Banff criteria.^14^ Each biopsy was considered a unique sample. As a result, some patients contributed multiple samples to our study: one patient contributed two samples to the non-rejection group, one patient contributed one rejection and one non-rejection sample, one patient contributed two rejection samples. The remaining 14 samples were obtained from unique patients.

#### CfDNA isolation

Blood (10 mL) was collected in EDTA tubes and stored at 4 degrees for no more than 24 hours. It was centrifuged at 2500g for 10 min at 4 degrees Celsius to separate plasma from cell components. Plasma was centrifuged a second time at 3500g for 10 additional minutes. Plasma was stored at -80 degrees Celsius until isolation.

cfDNA isolation was performed using the QIAMP CNA Kit, Catalogue number 55114 per manufacturer’s instructions. 3mL of plasma was first mixed with 300 μL of proteinase K and 2.4 mL buffer ACL, vortexed and incubated for 30 minutes at 60 degrees Celsius. Binding was optimized by the addition of 5.4 mL of Buffer ACB to lysate. Samples were then loaded onto the spin column using Qiagen vacuum manifold. They were washed first with 600 μL of ACW1 buffer, then 200 μL of ACW1 buffer, 750 μL of ACW2 buffer and 750 μL of 100% ethanol. The spin columns were transferred to a collection tube and spun at 20,000g for 3 minutes and incubated at 56 degrees Celsius for 10 minutes to dry the spin column of excess 100% ethanol. The DNA was eluted using 50 μL molecular-grade water and incubated for 10 minutes before centrifugation at 20,000g for 1 minute. Another elution was performed with 30 μL of molecular-grade water. CfDNA was then bead cleaned with AMPure XP Beads, Catalogue number A63881, with a 0.5x/1.6x size selection ratio. CfDNA was quantified using the Swift Input DNA Quantification Assay Protocol utilizing qPCR and stored at -20 degrees Celsius until library preparation.

#### Whole genome bisulfite sequencing (WGBS) library preparation and sequencing

A total of 5 ng of DNA was aliquoted from each sample. Unmethylated λDNA was added to each sample at 0.5% w/v. The samples were then concentrated with a SpeedVac to a volume of 20µL. Due to the fragmented nature of cfDNA, further fragmentation of the samples was not required. The samples then underwent bisulfite conversion with an EZ DNA Methylation-Gold kit (Zymo, Cat. No. D5006). The samples were eluted off the spin columns with 15μL of low EDTA TE buffer (Swift, Cat. No. 30024) before library preparation.

The low-input libraries were prepared using an ACCEL-NGS Methyl-Seq Library kit (Swift, Cat. No. 30024) with a Methyl-Seq Set A Indexing Kit (Swift, Cat. No. 36024), following the protocol associated with the library kit. During the protocol, bead cleanup steps were performed with SPRIselect beads (Beckman Coulter, Cat. No. B23318). Following the recommendation of the kit, 6 PCR cycles were performed to amplify the samples. The final libraries were quantified with a Qubit dsDNA HS Assay Kit and the size was determined by using a BioAnalyzer High Sensitivity DNA Kit (Agilent, Cat. No. 5067-4626). The libraries were then sequenced on the Illumina NovaSeq6000 System using 150bp paired-end sequencing.

#### WGBS data processing

WGBS data was processed using the Epigenome Pipeline available from the DRAGEN Bio-IT platform (Edico Genomics/Illumina). Sequence reads were demultiplexed into FASTQ files using Illumina’s bcl2Fastq2-2.19.1 software and trimmed for quality (phred33 >= 20) and Illumina adapters using trimgalore v.0.4.2 (https://github.com/FelixKrueger/TrimGalore). Reads were then aligned to the bisulfite-converted GRCh38 reference genome using DRAGEN EP v2.6.3 in paired-end mode using the directional/Lister methylation protocol presets. Alignments were calculated for both Watson and Crick strands and the highest quality unique alignment was retained. Duplicated reads were removed using picard v 2.17.8.^15^ A genome-wide cytosine methylation report was generated by DRAGEN to record counts of methylated and unmethylated cytosines at each cytosine position in the genome. Methylation counts were provided for the CpG, CHG and CHH cytosine contexts but only CpG was considered in the study. To avoid potential biases in downstream analyses, CpGs were further filtered by removing CpGs covered by five or less reads, and located within genomic regions that are known to have anomalous, unstructured, high signal/read counts as reported in DAC blacklisted regions (DBRs) or Duke excluded regions (DERs) generated by the ENCODE project.^16^

#### Differential methylation analysis

Filtered methylation data from all samples were merged according to histopathologic phenotype: non-rejection, borderline rejection, or rejection. Subsequently, CpGs were further filtered for final analysis, such that only those CpG sites covered by at least 10 reads and present in at least 50% of samples from the smaller of the comparison groups were included in analysis (i.e., present in ≥2 samples for comparisons against borderline rejection; present in ≥4 samples for comparisons of AAR vs non-rejection). DMCs of destranded autosomes were evaluated through a logistic regression analysis with time since transplant, recipient age, recipient/donor sex pairing (Female/Female, Female/Male, Male/Male, Male/Female), recipient race, and recipient WBC count included as covariates using the R-package, methylKit.^17^ *P-*values were adjusted to *q*-values using SLIM method. DMCs were defined as those with a *q*-value <0.05 and an absolute average methylation difference of >20% between comparison groups.

#### Gene Annotation

Following differential methylation analysis, gene annotation was performed on DMCs using Genomic Regions Enrichment of Annotations Tool (GREAT) algorithm ^18,19^ (Association rule: Single nearest gene: 5000 bp max extension, curated regulatory domains included) with identification of the single nearest gene regulatory domain within 5 kb of the DMC of interest. Subsequent gene ontology was further explored using Metascape^26^ with hypo- and hypermethylated DMCs evaluated separately.

#### Methylation segmentation

UMRs and LMRs for each samples set were called based on a pooled aggregate methylation profile across the samples using MethylSeekR package (v 1.38)^20^ from Bioconductor (v 3.16).^21,22^

MethylSeekR: Burger L, Gaidatzis D, Schubeler D, Stadler MB (2013). “Identification of active regulatory regions from DNA methylation data.” Nucleic Acids Research.

Doi:10.1093/nar/gkt599, http://nar.oxfordjournals.org/content/early/2013/07/04/nar.gkt599.long. Bioconductor: http://www.nature.com/nmeth/journal/v12/n2/abs/nmeth.3252.html, https://genomebiology.biomedcentral.com/articles/10.1186/gb-2004-5-10-r80

#### Annotation of regulatory elements

Genomic regions were further annotated for overlaps with the entire DNase I Hypersensitive Site (DHS) vocabulary using the *intersect* function in the Bedtools suite (v 2.30.0) ^23^ with minimum overlap of 1 nucleotide. The DHS coordinates were accessed from https://zenodo.org/record/3838751/files/DHS_Index_and_Vocabulary_hg38_WM20190703.txt.g z using 16 different vocabulary representatives as outlined in Meuleman *et al.*, 2020.^24^

BedTools: Quinlan AR and Hall IM, 2010. BEDTools: a flexible suite of utilities for comparing genomic features. Bioinformatics. 26, 6, pp. 841–842.

#### Transcription Factor Binding Analysis

Transcription factor binding site (TFBS) motif analysis was performed using the Homer software (HOMER findMotifsGenome.pl v4.11.1) ^20^ using the central 200bp of regions. Motif analysis was performed using HOMER software examining hypo- and hypermethylated DMCs separately, as well as looking at all DMCs (hypo- and hypermethylated) in aggregate.

## Results

### Characterization of plasma cfDNA methylome landscape by WGBS

From March 2019 to February 2023, 20 plasma samples were collected from pediatric kidney transplant recipients at the time of clinically indicated kidney biopsy (surveillance or for-cause) and cfDNA was isolated. Demographic features, underlying disease, and Banff classification^14^ of biopsy specimens are provided in Table 1.

**Table 1.**
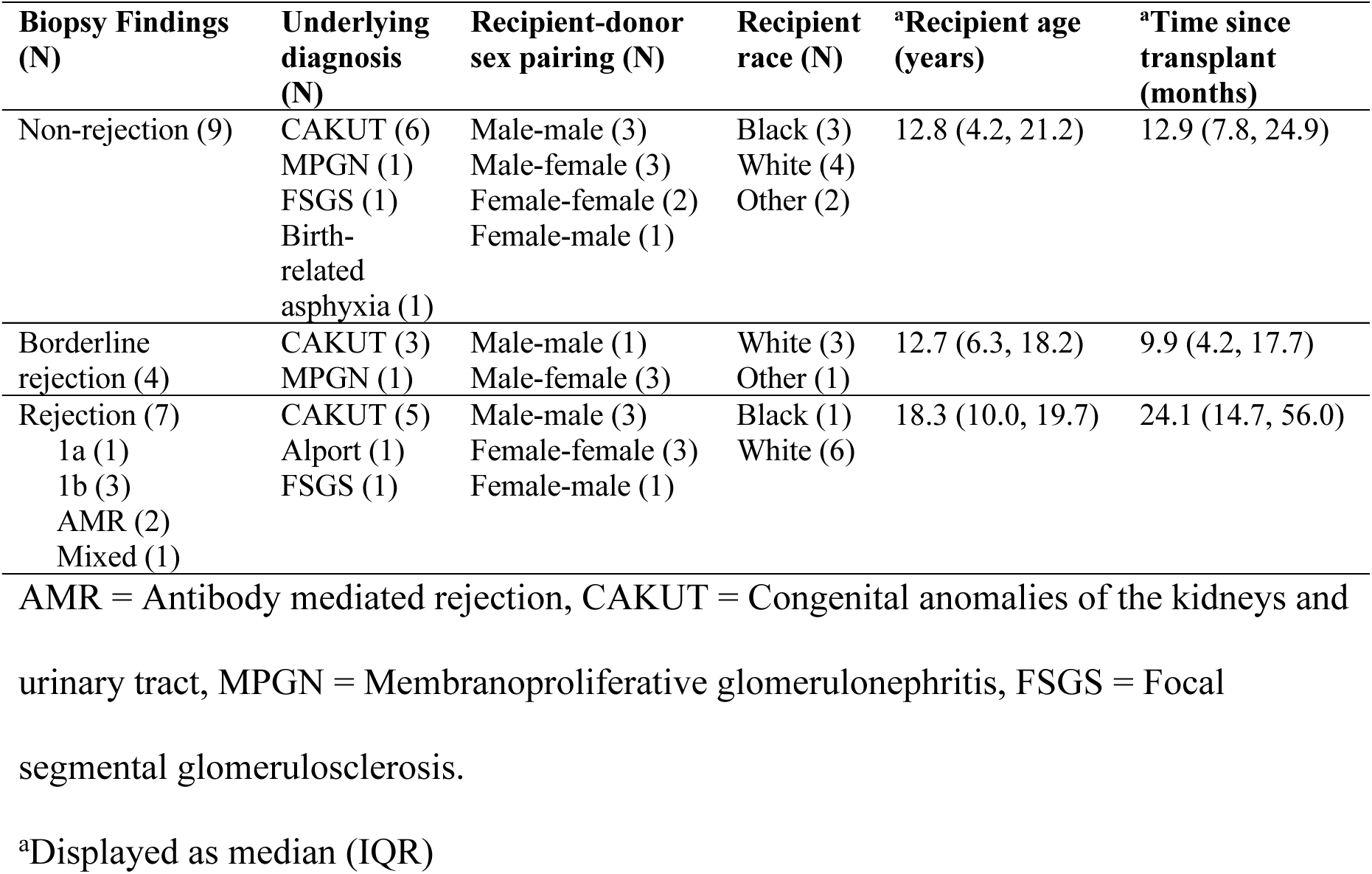
Banff classification of kidney biopsies,^14^ underlying diagnoses, and demographic features of subjects.

Whole genome bisulfite sequencing (WGBS) data was generated at high depth identifying on average 1.1 million CpGs per sample. Hierarchical clustering was performed on the most variably methylated residues (top 25^th^ percentile) of non-rejection (N = 9), borderline rejection (N = 4), and AAR (N = 7) groups and revealed no clustering structure (Supplemental Figure 1A). Hierarchical clustering was similarly performed on the most variably methylated residues (top 25^th^ percentile) for the categorical confounders of self-reported race and sex pairing of recipient and donor (*i.e.*, female-female, female-male, male-male, male-female) and no clustering structure was present (Supplemental Figure 1B, C).

Differentially methylated CpG residues (DMCs), defined as ≥20% difference in methylation rate and *q*-value <0.05, were compared between categories: borderline rejection vs non-rejection, AAR vs borderline rejection, and AAR vs non-rejection. These demonstrated 4,830 DMCs (2,683 hypomethylated, 2,147 hypermethylated); 3,536 DMCs (1,677 hypomethylated, 1,859 hypermethylated); and 34,365 DMCs (19,074 hypomethylated, 15,291 hypermethylated), respectively. We queried genes associated with DMCs using the Genomic Regions Enrichment of Annotations Tool (GREAT) algorithm.^18^ Comparison between the two most extreme phenotypes (AAR vs non-rejection) resulted in substantially higher counts of DMCs and their associated differentially methylated genes relative to comparisons made against borderline rejection samples. Specifically, 158 differentially methylated genes (86 hypo- and 75 hypermethylated) were noted between borderline rejection vs non-rejection groups; 98 differentially methylated genes (44 hypo- and 55 hypermethylated) between AAR vs borderline rejection groups; and 1,269 differentially methylated genes (718 hypo- and 593 hypermethylated) between AAR vs non-rejection groups (Table 2).

**Table 2.** Counts of differentially methylated cytosine residues (DMC), associated genes, and enriched biological pathways between borderline vs non-rejection, AAR vs borderline, and AAR vs non-rejection groups. Note that total genes and biological pathways represent unique values and are less than the sums of hypo- and hypermethylated counts, as some genes and pathways include both hypo- and hypermethylated DMCs.

| **Comparison groups** | **DMC Count** | **Genes** | **Biological Pathways** |
| --- | --- | --- | --- |
| Borderline vs Non-rejection | Total: 4,830<br>Hypomethylated: 2,683<br>Hypermethylated: 2,147 | Total: 158<br>Hypomethylated: 86<br>Hypermethylated: 75 | Total: 128<br>Hypomethylated: 70<br>Hypermethylated: 72 |
| AAR vs Borderline | Total: 3,536<br>Hypomethylated: 1,677<br>Hypermethylated: 1,859 | Total: 98<br>Hypomethylated: 44<br>Hypermethylated: 55 | Total: 110<br>Hypomethylated: 36<br>Hypermethylated: 86 |
| AAR vs Non-rejection | Total: 34,365<br>Hypomethylated: 19,074<br>Hypermethylated: 15,291 | Total: 1,269<br>Hypomethylated: 718<br>Hypermethylated: 593 | Total: 533<br>Hypomethylated: 345<br>Hypermethylated: 222 |

We then characterized active regulatory regions of the plasma cfDNA through methylation segmentation to extract unmethylated regions (UMRs) and low methylated regions (LMRs), which correlate with promoter- and enhancer-like elements, respectively.^25^ Aggregating the samples, we identified 17,002 UMRs (average 2,212 bp) with DNA methylation levels across the regions being <5% and containing on average 34 CpGs per region. LMRs were more abundant, as we identified 46,265 regions with an intermediate methylation status (5-50%) which were more CpG-sparse (an average of 6 CpGs per region with an average 575 bp in length) than UMRs. We annotated UMRs and LMRs to publicly available regulatory DNA reference maps based on DNase I hypersensitive sites (DHSs) across 16 different cell types^24^ and found 99% and 98% of UMRs and LMRs, respectively, overlapped a DHS. Of these annotated regions, the vast majority (98%) of UMRs overlapped a DHS detected in multiple cell types, in-line with known epigenomic features of promoter regions. Relative to UMRs, LMRs were shown to represent to a larger extent cell-specific regulatory DNA, with 31% of LMRs (N= 14,407) overlapping a DHS unique to a specific cell type. Of these 14,407 LMRs we noted 11.5% and 30.1% being lymphoid and myeloid regulatory elements, respectively. Apart from borderline samples demonstrating relatively higher proportions of myeloid regulatory elements (34.7%, *p* <0.001) and lower proportion of embryonic primitive regulatory elements (1.0%, *p* <0.001), proportions of other cell type-specific DHSs were equivalent among the three comparisons (Table 3).

**Table 3.**
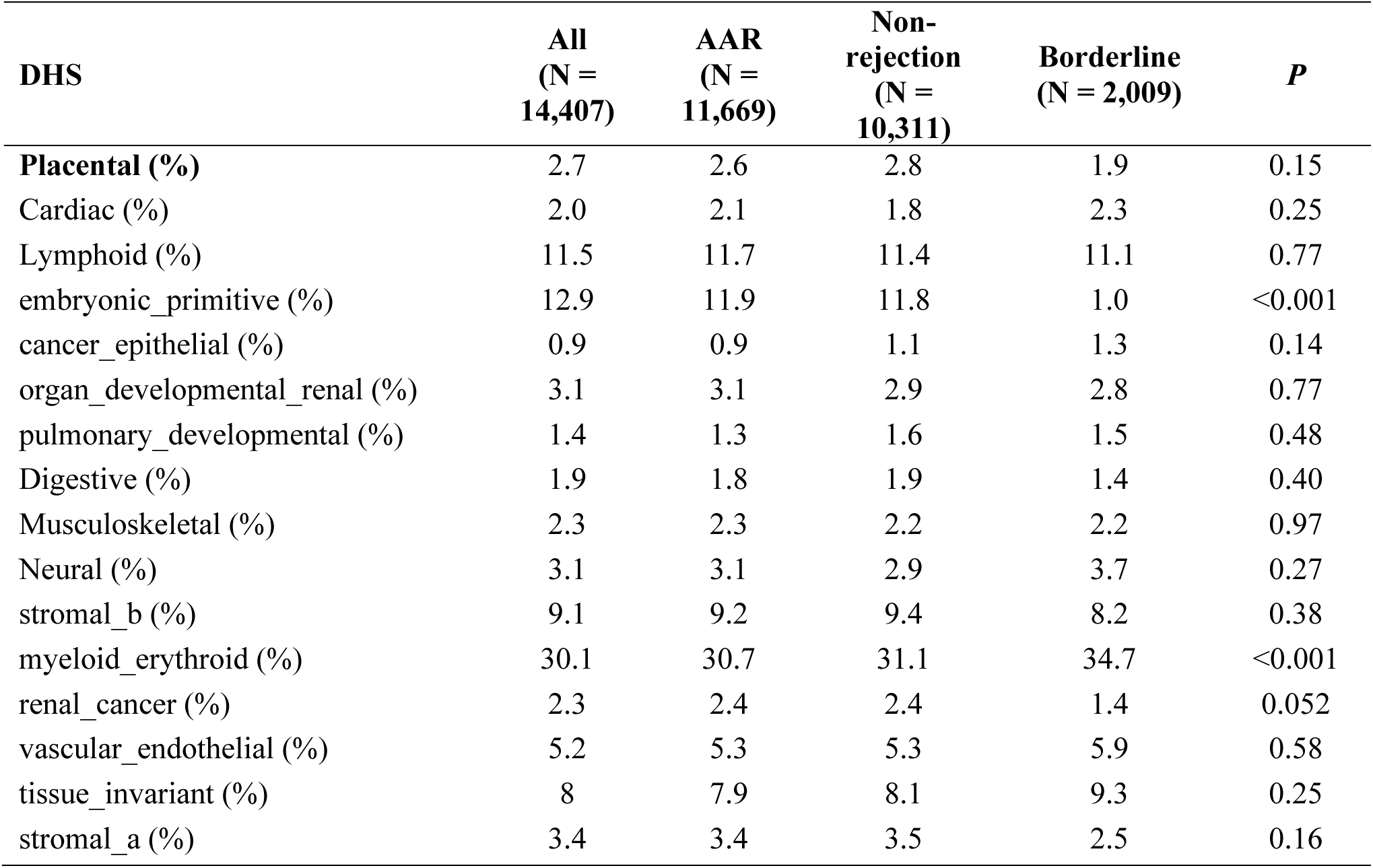
Annotation of UMRs and LMRs across all sampled individuals, AAR, non-rejection, and borderline rejection, to regulatory DNA reference maps^24^ based on DNase I hypersensitive sites (DHSs) across 16 different cell types.

| **DHS** | **All**<br>**(N =**<br>**14,407)** | **AAR**<br>**(N =**<br>**11,669)** | **Non-**<br>**rejection**<br>**(N =**<br>**10,311)** | **Borderline**<br>**(N = 2,009)** | ***P*** |
| --- | --- | --- | --- | --- | --- |
| **Placental (%)** | 2.7 | 2.6 | 2.8 | 1.9 | 0.15 |
| Cardiac (%) | 2.0 | 2.1 | 1.8 | 2.3 | 0.25 |
| Lymphoid (%) | 11.5 | 11.7 | 11.4 | 11.1 | 0.77 |
| embryonic_primitive (%) | 12.9 | 11.9 | 11.8 | 1.0 | <0.001 |
| cancer_epithelial (%) | 0.9 | 0.9 | 1.1 | 1.3 | 0.14 |
| organ_developmental_renal (%) | 3.1 | 3.1 | 2.9 | 2.8 | 0.77 |
| pulmonary_developmental (%) | 1.4 | 1.3 | 1.6 | 1.5 | 0.48 |
| Digestive (%) | 1.9 | 1.8 | 1.9 | 1.4 | 0.40 |
| Musculoskeletal (%) | 2.3 | 2.3 | 2.2 | 2.2 | 0.97 |
| Neural (%) | 3.1 | 3.1 | 2.9 | 3.7 | 0.27 |
| stromal_b (%) | 9.1 | 9.2 | 9.4 | 8.2 | 0.38 |
| myeloid_erythroid (%) | 30.1 | 30.7 | 31.1 | 34.7 | <0.001 |
| renal_cancer (%) | 2.3 | 2.4 | 2.4 | 1.4 | 0.052 |
| vascular_endothelial (%) | 5.2 | 5.3 | 5.3 | 5.9 | 0.58 |
| tissue_invariant (%) | 8 | 7.9 | 8.1 | 9.3 | 0.25 |
| stromal_a (%) | 3.4 | 3.4 | 3.5 | 2.5 | 0.16 |

### Differentially methylated cytosines predominately lie in intergenic and intronic regions

Given the much more striking difference between AAR and non-rejection groups relative to comparisons against the borderline rejection group, the remainder of our analyses principally focused on comparisons of AAR vs non-rejection. In the comparison of DMCs between AAR vs non-rejection, the majority (52.3%) of cytosine residues localized to intergenic regions, 44.3% to introns, and Σ1% each to triplex target DNA sites (TTS), 3’ untranslated regions (UTR), exons, promoter – transcription start site (TSS), non-coding regions, and 5’ UTR (Supplemental Table 1).

### Genes related to lipid metabolism, tryptophan-kynurenine metabolism, inflammatory **response, and immune cell regulation are differentially methylated in AAR**

To determine differentially methylated pathways in relation to the differentially methylated genes noted between comparison groups, pathway enrichment analysis was performed using Metascape^26^ with a *p*-value threshold of ≤0.01 and found 345 and 222 enriched hypo- and hypermethylated pathways, respectively (Supplemental Table 2A, B). Emergent themes identified in both hypo- and hypermethylated analyses included processes involved in immune cell regulation, inflammatory response, and lipid metabolism (Figure 1A, B). Additionally, pathways associated with tryptophan-kynurenine metabolism were enriched in hypermethylated genes (Figure 1B).

**Figure 1.**
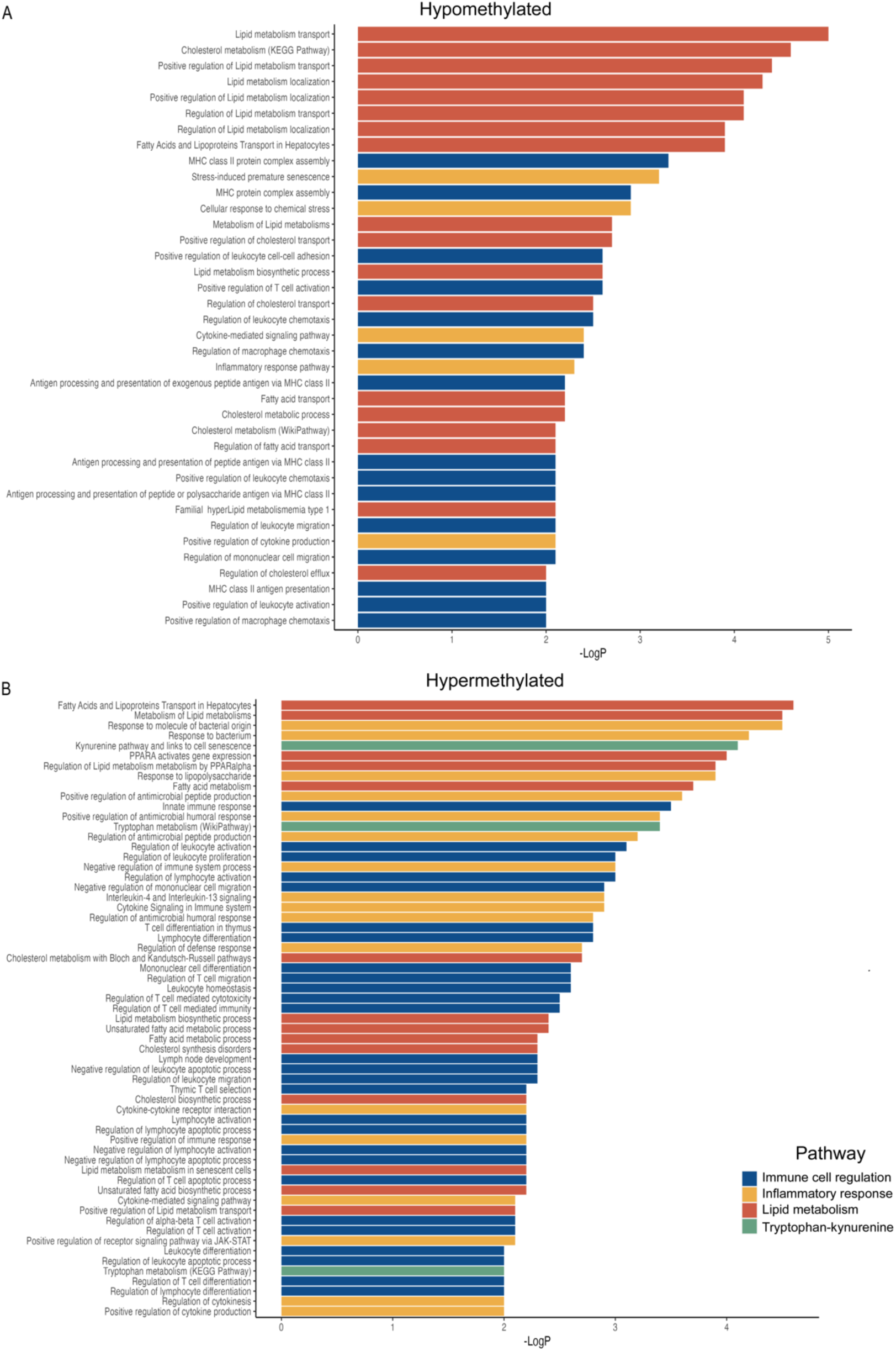
Enriched biological pathways of (A) hypomethylated and (B) hypermethylated plasma cfDNA of individuals with AAR vs non-rejection demonstrate differential regulation of pathways related to immune cell regulation (blue), inflammatory response (yellow), lipid metabolism (orange), and tryptophan-kynurenine metabolism (green). Enrichment analysis was carried out using Metascape^26^ and included Gene Ontology (GO) terms,^50^ Kyoto Encyclopedia of Genes and Genomes (KEGG) pathways,^51^ Reactome gene sets,^52^ and WikiPathways^53^ (y-axis). The -log_10_P values is represented on the x-axis.

Notably, there were multiple enriched pathways associated with lipid metabolism in the comparison of borderline vs non-rejection. Predominate themes from enriched pathways between AAR and borderline groups included lipid metabolism, small molecule transport, and cell transport (Supplemental Tables 2C-F).

### Differential methylation of innate and adaptive immune cell populations contributes to **AAR pathophysiology**

A total of 101 differentially methylated genes from our dataset function in pathways related to immune cell regulation in innate and/or adaptive immune response. Of these, 29 were hypomethylated and 74 hypermethylated in ARR relative to non-rejection (Supplemental Table 3), with *APOD* and *SHH* containing both hypo- and hypermethylated DMCs at different loci (Table 4, Figure 2).

**Table 4.**
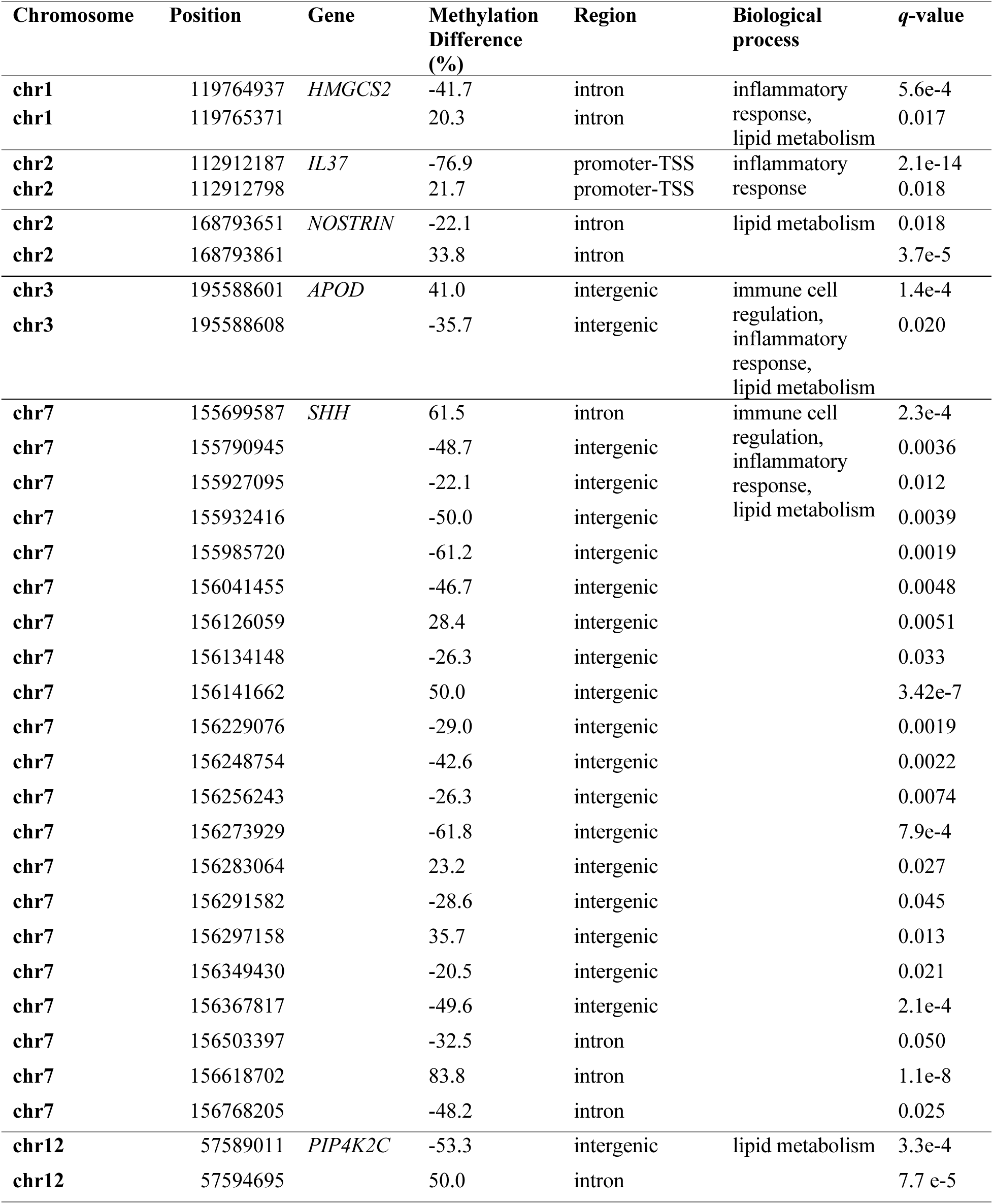
Genes containing relatively hypomethylated and hypermethylated DMCs in AAR vs non-rejection. Negative methylation difference values indicate hypomethylation.

**Figure 2.**
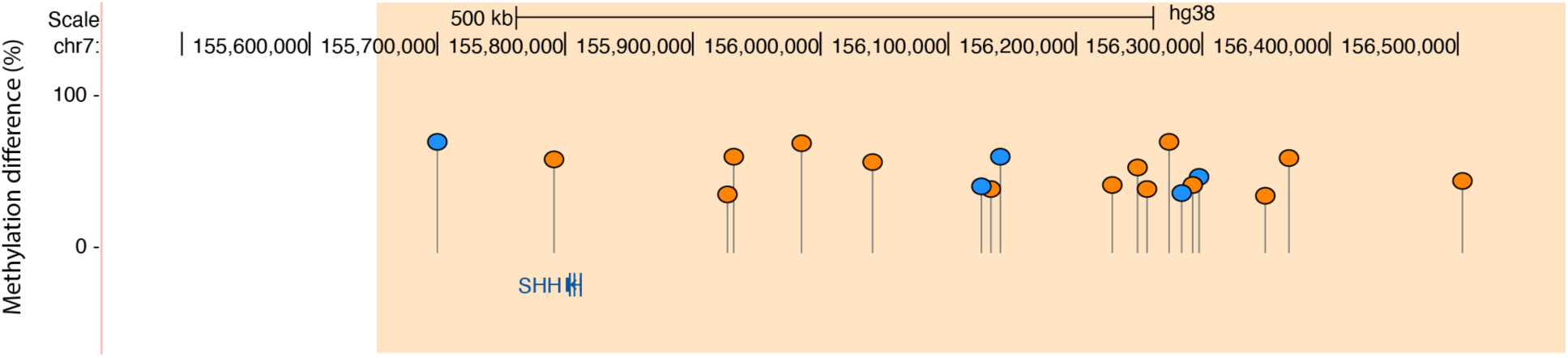
DMCs in AAR vs non-rejection associated with *SHH* curated regulatory domain (chr7: 155645508-156791875) as annotated by GREAT algorithm.^18^. Y-axis demonstrates percent methylation difference of DMCs between AAR and non-rejection. Relatively hypomethylated DMCs are present in orange; relatively hypermethylated in blue. The area falling within the *SHH* curated domain is indicated in the tan rectangle.

Genes for which there is particularly abundant evidence in the literature relating them to transplantation included *FASLG*, *IRF8*, *NLRP2*, *IL17F*, *IL7R*, and *IRF4* which contained hypermethylated DMCs; and *AIF1*, *NLRP3*, and *HMGB1* which contained hypomethylated DMCs.^27–39^ We also noted multiple biological pathways associated with major histocompatibility complex (MHC) class II, including hypomethylation in *HLA-DOA*, *HLA-DQA1*, and *HLA-DQB2*.

Notable hypomethylation in AAR was present in *SHH*, including a single DMC (chr7:155790945, methylation difference = -48%, *q* = 0.003) located downstream of the coding region and 14 hypomethylated DMCs residing upstream of coding and promoter regions (chr7: 155927095-156768205, methylation difference range = -22% to -61%, *q*-value range = 7.95 x 10^-4^ to 0.05). Similarly, a single hypermethylated DMC associated with *SHH* was present downstream of the coding region (chr7: 155699587, methylation difference = +61%, *q* = 2.32 x 10^-4^), while an additional five DMCs were located upstream of coding and promoter regions (chr7: 156126059-156618702, methylation difference = +35 to +83%, *q*-value range = 0.027 – 1.07 x 10^-8^) (Figure 2, Table 4).

There were 142 differentially methylated genes associated with biological pathways related to the inflammatory response, of which 45 contained relatively hypomethylated DMCs and 98 contained relatively hypermethylated DMCs in AAR vs non-rejection (*IL37* contained hypo- and hypermethylated DMCs) (Supplemental Table 4). There was substantial overlap in differentially methylated genes between pathways involved in immune cell regulation and the inflammatory response, with 66 shared differentially methylated genes (Supplemental Tables 3, 4). Notably, *PCNA* (chr20:5125534, methylation difference = -44%, *q* = 7.44 x 10^-4^) and *P2RX7* (chr12:121131322, methylation difference = -45%, *q* = 0.02) contained hypomethylated DMCs.

Cumulatively, these findings suggest cytokines and the inflammatory response work to modulate the activity and expansion of lymphocytes in their contribution to rejection pathophysiology and highlight the role of innate immune cells in AAR pathogenesis. Further, these findings suggest differential methylation status of human leukocyte antigen (HLA) genes contribute to rejection risk.

### Lipid metabolism modulates inflammatory pathways

Comparing differentially methylated pathways between AAR vs non-rejection, we found 32 pathways associated with lipid metabolism (17 hypomethylated, 15 hypermethylated) with 125 differentially methylated genes (72 hypomethylated, 57 hypermethylated, with *HMGCS2*, *SHH*, *NOSTRIN*, and *PIP4K2C* containing hypo- and hypermethylated DMCs) (Supplemental Table 5, Table 4). Notable differentially methylated genes within these pathways included hypomethylation of *P2RX7*, *APOL1*, *APOL3*, *APOL5*, and *SIRT1*; and hypermethylation of *PIK3C3*. These data indicate that the inflammatory response may be modulated by interactions with lipid metabolism pathways, and that differential methylation plays a contributory role to these processes.

### Tryptophan-kynurenine pathway metabolites modulate cytokine expression in allograft **rejection**

In our comparison of AAR vs non-rejection samples, three hypermethylated biological pathways related to the tryptophan-kynurenine pathway were noted with seven associated relatively hypermethylated genes (*AHR*, *CDKN1A*, *IDO1*, *ACMSD*, *EIF2AK4*, *DDC*, and *TPH1*) (Supplemental Table 6).

These data suggest that interplay between the tryptophan-kynurenine pathway and cytokine response exists as a mediator of allograft rejection, with methylation contributing to the regulation of these pathways.

### Differential methylation of hypoxia inducible factors binding domains contributes to **rejection pathophysiology**

To gain further insights into the regulatory pathways involved in AAR pathogenesis, we examined DMCs within transcription factor binding sites (TFBS) in AAR vs non-rejection samples through HOMER^20^ motif analysis, with hypo- and hypermethylated DMCs once again evaluated separately (Supplemental Table 7A, B). This approach identified a significant enrichment of hypomethylation of TFBS for retinoic acid receptor-alpha (RARα), hypermethylation of TFBS for HIF-1α and HIF-2α, and hypo- and hypermethylation of TFBS for HIF-1ϕ3 and the ARNT:AhrR complex.

## Discussion

In our interrogation of the plasma cfDNA methylome in kidney transplant recipients, we highlight the importance of methylation in AAR pathophysiology. Many of our findings build upon prior knowledge of molecular mechanisms driving rejection, however, our novel approach of evaluating methylation status in the real-time biomarker, cfDNA, implicates methylation as a contributory player in these pathogenic pathways. Our demonstration of a large proportion of LMRs and intronic and intergenic regions among the DMCs in these samples suggests differential methylation in enhancer-type regions and distal regulatory elements are among the principal means by which methylation contributes to kidney allograft rejection.

Though AAR is typically viewed as a lymphocyte-mediated disease, we identified differential methylation of genes across many innate immune cells, including monocytes, PBMCs, and neutrophils. We used the Human Protein Atlas (https://www.proteinatlas.org)^40^ to discern which cell types within our dataset may have been impacted by the differentially methylated genes we observed in AAR vs non-rejection. Human Protein Atlas data indicated that *FASLG* and *IL7R* are predominately expressed in T-cell subsets. *IRF8* and *IRF4* are predominately expressed in dendritic cells (DCs); however, *IRF4* is also heavily expressed in naïve and memory B-cells with a lower level of expression in T-regulatory cells (Supplemental Figure 2A-D). *AIF1* is principally expressed in monocytes but also has expression in peripheral blood mononuclear cells (PBMCs), neutrophils, and myeloid DCs; *HLA-DOA* is expressed in B-cell subsets but also in myeloid DCs; *HLA-DQA1* expressed in DCs; *HLA-DQB2* is expressed in memory B-cells and DCs, albeit at low levels; *NLRP3* is highly expressed in basophils; and *HMGB1* is fairly non-specific in its immune cell specificity (Supplemental Figure 2E-J). This suggests that, in addition to T-cell and B-cell mediated processes, differential methylation of genes related to innate immunity contributes to AAR pathophysiology.

Our study suggests an interplay between lipid metabolism and the immune response. For example, we identified hypermethylation associated with the *PIK3C3* gene in those with AAR as compared to non-rejection. The phosphoinositide 3-kinase (PI3K) pathway has been implicated in acute rejection of solid organ allografts due to its role in promoting differential activation of CD4^+^ T cell subsets. More specifically, PIK3C3 is a component of the class III lipid kinase complex I (PI3KC3-C1).^41^ PI3KC3-C1 is essential for autophagy. Dysregulation of normal macroautophagy leads to impaired cell survival and stress responses in various human diseases.^42^ In this way, it appears that altered methylation of lipid metabolism pathways modulates immune cell function and survival as a component of rejection pathogenesis, or perhaps as a response to AAR.

We demonstrate differential methylation within the tryptophan-kynurenine pathway between individuals with AAR vs non-rejection, including differential methylation of *AHR*, *CDKN1A*, *IDO1*, *ACMSD*, *EIF2AK4*, *DDC*, and *TPH1*. Indoleamine-2,3-dioxygenase 1 (IDO1), the gene product of *IDO1*, is an early rate-limiting enzyme in the tryptophan-kynurenine pathway. IDO1 is expressed broadly across antigen presenting cells with increased expression in response to inflammation. IDO1 then acts to decrease tryptophan levels, which has been linked to T-cell anergy, T-cell cycle arrest, and apoptosis. A wealth of data has been produced using animal models linking IDO1 to decreased allograft rejection, induction of allograft tolerance, and improved graft survival across many solid organ transplant tissue types (kidney, heart, lung, pancreas, small bowel). Further downstream in the tryptophan-kynurenine pathway, inhibition of kynurenine-3-monooxygenase (KMO) (not among the differentially methylated genes in our dataset) has been associated with reduction in inflammation through activation of the aryl hydrocarbon receptor (AhR) gene product of *AHR* and secretion of the anti-inflammatory cytokine, interleukin-4 (IL-4), by natural killer cells.^43^ AhR expression has been linked to allograft tolerance in skin, pancreas, and cardiac transplants due to its role in reducing generation and proliferation of cytotoxic T lymphocytes, and also due to its modulation of the innate immune response through repression of acute phase reactants.^44^ Interestingly, hypermethylation of IL-4 and interleukin-13 (IL-13) signaling was among the many biological pathways related to the inflammatory and cytokine responses from our gene list (Figure 1B). The anti-inflammatory cytokine, interleukin-4 (IL-4) has been linked to the tryptophan-kynurenine pathway by the activation of the aryl hydrocarbon receptor (AhR) gene product of *AHR*, which acts to promote secretion of IL-4 by natural killer cells.^43^

With regard to our TFBS analysis, we identified binding sites of HIF-1α, HIF-1β, HIF-2α, and the ARNT:AhR complex to be significantly enriched. Hypoxia-inducible factors (HIFs) are transcription factors that function as heterodimers in an adaptive response to hypoxia and are composed of an alpha subunit controlled in an oxygen dependent fashion and a stably expressed beta subunit. The HIF-1β subunit, also known as aryl hydrocarbon receptor nuclear translocators (ARNT), is an obligate partner of the aryl hydrocarbon receptor (AHR).^45^ HIFs, particularly HIF-1α, have been shown to directly correlate with clinical and subclinical rejection in kidney allografts.^46,47^ This provides strong support to existing data indicating that HIFs and methylation statuses of their targets contribute to rejection pathophysiology and/or response to rejection in the kidney allograft. Hypomethylation of RARα targets were also appreciated. Prior studies demonstrate that RARα agonists prevent acute and chronic allograft rejection. These effects are enhanced by activation of peroxisome proliferators-activated receptor-ψ (PPARψ)^48,49^—note that *PPARG* contained a hypermethylated DMC in our comparison of AAR vs non-rejection (chr3:12349708, methylation difference = +45.18%, *q* = 3.0 x 10^-4^).

When viewed in aggregate, we see considerable overlap and interactions between pathways, transcription factors, and genes in the predominate biological pathways we highlighted. This suggests the processes of lipid metabolism, inflammation, immune cell regulation, and the tryptophan-kynurenine pathway do not act in isolation to mediate AAR. Rather, there is an interplay of these biological processes in the development of AAR or response to AAR, with differential methylation being a contributory factor.

There are some limitations to this study. Due the nature of this pilot study and because pediatric kidney transplant patients are limited in number relative to adult transplant recipients, our sample size was limited. This small sample size paired with the heterogeneity of underlying diagnoses in our sample population could have led to interindividual methylation differences unrelated to rejection status. To overcome this potential for encountering incidental methylation differences, we set a conservative threshold for defining significant DMCs (>20% methylation difference, *q* <0.05). However, this comes with the inherent consequence of excluding potentially important DMCs that contribute to rejection pathophysiology. Additionally, our coverage of the genome is limited to approximately 4% of total CpG sites. This likely comes from the combination of our small sample size and the degradation of already fragmented cfDNA through sodium bisulfite conversion. Finally, given that samples were collected at time of biopsy, it is difficult to ascertain whether our findings reflect drivers of rejection pathogenesis or reactions to rejection.

In conclusion, differential methylation in pathways associated with inflammation, immune cell regulation, lipid metabolism, and tryptophan-kynurenine metabolism appear to play a role in the pathophysiology of acute allograft rejection in pediatric kidney transplantation. Though further studies with more robust sample sizes are needed, our findings emphasize the need to examine epigenetics in the context of allograft health and dysfunction in solid organ transplantation.

## Data Availability

Raw and processed data are available via the Gene Expression Omnibus (WGBS accession number: GSE275017, URL: https://www.ncbi.nlm.nih.gov/geo/query/acc.cgi?acc=GSE275017

## Acknowledgements

The authors would like to thank Margaret Gibson and Rebecca Biswell for their work in sample processing, Bradley Belden and Macy Mcbeth for their roles in patient recruitment and sample collection, and the Children’s Mercy Hospital Transplant Coordinators for their assistance in subject identification. BLS was supported in part by The Sam and Helen Kaplan Research Fund in Pediatric Nephrology, and The McLaughlin Family Endowed Chair in Nephrology. This research was presented in part in oral format at the American Society of Nephrology Kidney Week 2023 and as a poster presentation at the Pediatric Academic Societies 2024 annual meeting.

## Data Sharing Statement

Raw and processed data are available via the Gene Expression Omnibus (WGBS accession number: GSE275017, URL: https://www.ncbi.nlm.nih.gov/geo/query/acc.cgi?acc=GSE275017).

## References

1. System USRD. 2019 *USRDS Annual Data Report: Epidemiology of kidney disease in the United States*. 2019. https://www.usrds.org/annual-data-report/

2. Poggio ED, Augustine JJ, Arrigain S, Brennan DC, Schold JD. Long-term kidney transplant graft survival-Making progress when most needed. Am J Transplant. Aug 2021;21(8):2824–2832. doi:10.1111/ajt.16463

3. Lamb KE, Lodhi S, Meier-Kriesche HU. Long-term renal allograft survival in the United States: a critical reappraisal. Am J Transplant. Mar 2011;11(3):450–462. doi:10.1111/j.1600-6143.2010.03283.x

4. Clayton PA, McDonald SP, Russ GR, Chadban SJ. Long-Term Outcomes after Acute Rejection in Kidney Transplant Recipients: An ANZDATA Analysis. J Am Soc Nephrol. Sep 2019;30(9):1697–1707. doi:10.1681/asn.2018111101

5. Owoyemi I, Tandukar S, Jorgensen DR, et al. Impact of Subclinical and Clinical Kidney Allograft Rejection Within 1 Year Posttransplantation Among Compatible Transplant With Steroid Withdrawal Protocol. Transplantation Direct. 2021;7(7):e706. doi:10.1097/txd.0000000000001132

6. El-Zoghby ZM, Stegall MD, Lager DJ, et al. Identifying specific causes of kidney allograft loss. Am J Transplant. Mar 2009;9(3):527–535. doi:10.1111/j.1600-6143.2008.02519.x

7. Spector BL, Harrell L, Sante D, Wyckoff GJ, Willig L. The methylome and cell-free DNA: current applications in medicine and pediatric disease. Pediatr Res. Jul 2023;94(1):89–95. doi:10.1038/s41390-022-02448-3

8. Xiao H, Gao F, Pang Q, et al. Diagnostic Accuracy of Donor-derived Cell-free DNA in Renal-allograft Rejection: A Meta-analysis. Transplantation. 2021;105(6)

9. Bloom RD, Bromberg JS, Poggio ED, et al. Cell-Free DNA and Active Rejection in Kidney Allografts. J Am Soc Nephrol. Jul 2017;28(7):2221–2232. doi:10.1681/asn.2016091034

10. Huang E, Sethi S, Peng A, et al. Early clinical experience using donor-derived cell-free DNA to detect rejection in kidney transplant recipients. Am J Transplant. Jun 2019;19(6):1663–1670. doi:10.1111/ajt.15289

11. Witasp A, Luttropp K, Qureshi AR, et al. Longitudinal genome-wide DNA methylation changes in response to kidney failure replacement therapy. Sci Rep. Jan 10 2022;12(1):470. doi:10.1038/s41598-021-04321-5

12. Mas VR, Le TH, Maluf DG. Epigenetics in Kidney Transplantation: Current Evidence, Predictions, and Future Research Directions. Transplantation. Jan 2016;100(1):23–38. doi:10.1097/tp.0000000000000878

13. Zhu C, Xiang W, Li B, et al. DNA methylation modulates allograft survival and acute rejection after renal transplantation by regulating the mTOR pathway. (1600-6135 (Print))

14. Haas M, Loupy A, Lefaucheur C, et al. The Banff 2017 Kidney Meeting Report: Revised diagnostic criteria for chronic active T cell-mediated rejection, antibody-mediated rejection, and prospects for integrative endpoints for next-generation clinical trials. Am J Transplant. Feb 2018;18(2):293–307. doi:10.1111/ajt.14625

15. Broad Institute. Picard toolkit. 2019. http://broadinstitute.github.io/picard/

16. Amemiya HM, Kundaje A, Boyle AP. The ENCODE Blacklist: Identification of Problematic Regions of the Genome. Scientific Reports. 2019/06/27 2019;9(1):9354. doi:10.1038/s41598-019-45839-z

17. Akalin A, Kormaksson M, Li S, et al. methylKit: a comprehensive R package for the analysis of genome-wide DNA methylation profiles. Genome Biology. 2012/10/03 2012;13(10):R87. doi:10.1186/gb-2012-13-10-r87

18. McLean CY, Bristor D, Hiller M, et al. GREAT improves functional interpretation of cis-regulatory regions. Nat Biotechnol. May 2010;28(5):495–501. doi:10.1038/nbt.1630

19. Tanigawa Y, Dyer ES, Bejerano G. WhichTF is functionally important in your open chromatin data? PLoS Comput Biol. Aug 2022;18(8):e1010378. doi:10.1371/journal.pcbi.1010378

20. Burger L, Gaidatzis D, Schübeler D, Stadler MB. Identification of active regulatory regions from DNA methylation data. Nucleic Acids Research. 2013;41(16):e155–e155. doi:10.1093/nar/gkt599

21. Huber W, Carey VJ, Gentleman R, et al. Orchestrating high-throughput genomic analysis with Bioconductor. Nature Methods. 2015/02/01 2015;12(2):115–121. doi:10.1038/nmeth.3252

22. Gentleman RC, Carey VJ, Bates DM, et al. Bioconductor: open software development for computational biology and bioinformatics. Genome Biology. 2004/09/15 2004;5(10):R80. doi:10.1186/gb-2004-5-10-r80

23. Quinlan AR, Hall IM. BEDTools: a flexible suite of utilities for comparing genomic features. Bioinformatics. Mar 15 2010;26(6):841–842. doi:10.1093/bioinformatics/btq033

24. Meuleman W, Muratov A, Rynes E, et al. Index and biological spectrum of human DNase I hypersensitive sites. Nature. Aug 2020;584(7820):244–251. doi:10.1038/s41586-020-2559-3

25. Busche S, Shao X, Caron M, et al. Population whole-genome bisulfite sequencing across two tissues highlights the environment as the principal source of human methylome variation. Genome Biol. Dec 23 2015;16:290. doi:10.1186/s13059-015-0856-1

26. Zhou Y, Zhou B, Pache L, et al. Metascape provides a biologist-oriented resource for the analysis of systems-level datasets. Nat Commun. Apr 3 2019;10(1):1523. doi:10.1038/s41467-019-09234-6

27. Heng B, Ding H, Ren H, et al. Diagnostic Performance of Fas Ligand mRNA Expression for Acute Rejection after Kidney Transplantation: A Systematic Review and Meta-Analysis. PLoS One. 2016;11(11):e0165628. doi:10.1371/journal.pone.0165628

28. Franchon Marques Tejada N, Ziroldo Lopes JV, Duarte Gonçalves LE, et al. AIM2 as a putative target in acute kidney graft rejection. Front Immunol. 2022;13:839359. doi:10.3389/fimmu.2022.839359

29. Shen Q, Wang Y, Chen J, et al. Single-Cell RNA Sequencing Reveals the Immunological Profiles of Renal Allograft Rejection in Mice. Front Immunol. 2021;12:693608. doi:10.3389/fimmu.2021.693608

30. Granell M, Urbano-Ispizua A, Pons A, et al. Common variants in NLRP2 and NLRP3 genes are strong prognostic factors for the outcome of HLA-identical sibling allogeneic stem cell transplantation. Blood. Nov 15 2008;112(10):4337–4342. doi:10.1182/blood-2007-12-129247

31. Haouami Y, Dhaouadi T, Sfar I, et al. The role of IL-23/IL-17 axis in human kidney allograft rejection. J Leukoc Biol. Dec 2018;104(6):1229–1239. doi:10.1002/jlb.5ab0318-148r

32. Gorbacheva V, Fan R, Li X, Valujskikh A. Interleukin-17 promotes early allograft inflammation. Am J Pathol. Sep 2010;177(3):1265–1273. doi:10.2353/ajpath.2010.091106

33. Mai HL, Boeffard F, Longis J, et al. IL-7 receptor blockade following T cell depletion promotes long-term allograft survival. J Clin Invest. Apr 2014;124(4):1723–1733. doi:10.1172/jci66287

34. Schreiber M, Weigelt M, Karasinsky A, et al. Inducible IL-7 Hyperexpression Influences Lymphocyte Homeostasis and Function and Increases Allograft Rejection. Front Immunol. 2019;10:742. doi:10.3389/fimmu.2019.00742

35. Wu J, Zhang H, Shi X, et al. Ablation of Transcription Factor IRF4 Promotes Transplant Acceptance by Driving Allogenic CD4(+) T Cell Dysfunction. Immunity. Dec 19 2017;47(6):1114–1128.e1116. doi:10.1016/j.immuni.2017.11.003

36. Zou D, Fu J, Guo Z, Chen W. Interferon regulatory factor 4 deficiency in CD8(+) T cells abrogates terminal effector differentiation and promotes transplant acceptance. Immunology. Dec 2020;161(4):364–379. doi:10.1111/imm.13258

37. Sikora M, Kopeć B, Piotrowska K, Pawlik A. Role of allograft inflammatory factor-1 in pathogenesis of diseases. Immunol Lett. Feb 2020;218:1–4. doi:10.1016/j.imlet.2019.12.002

38. Huang Y, Yin H, Han J, et al. Extracellular hmgb1 functions as an innate immune-mediator implicated in murine cardiac allograft acute rejection. Am J Transplant. Apr 2007;7(4):799–808. doi:10.1111/j.1600-6143.2007.01734.x

39. Chen Y, Zhang W, Bao H, He W, Chen L. High Mobility Group Box 1 Contributes to the Acute Rejection of Liver Allografts by Activating Dendritic Cells. Front Immunol. 2021;12:679398. doi:10.3389/fimmu.2021.679398

40. Uhlén M, Fagerberg L, Hallström BM, et al. Proteomics. Tissue-based map of the human proteome. Science. Jan 23 2015;347(6220):1260419. doi:10.1126/science.1260419

41. Lu G, Wang Y, Shi Y, et al. Autophagy in health and disease: From molecular mechanisms to therapeutic target. MedComm (2020). Sep 2022;3(3):e150. doi:10.1002/mco2.150

42. Xie Y, Li J, Kang R, Tang D. Interplay Between Lipid Metabolism and Autophagy. Front Cell Dev Biol. 2020;8:431. doi:10.3389/fcell.2020.00431

43. Zulpaite R, Miknevicius P, Leber B, Strupas K, Stiegler P, Schemmer P. Tryptophan Metabolism via Kynurenine Pathway: Role in Solid Organ Transplantation. Int J Mol Sci. Feb 15 2021;22(4)doi:10.3390/ijms22041921

44. Gargaro M, Pirro M, Romani R, Zelante T, Fallarino F. Aryl Hydrocarbon Receptor– Dependent Pathways in Immune Regulation. American Journal of Transplantation. 2016/08/01/ 2016;16(8):2270–2276. 10.1111/ajt.13716

45. Dengler VL, Galbraith M, Espinosa JM. Transcriptional regulation by hypoxia inducible factors. Crit Rev Biochem Mol Biol. Jan-Feb 2014;49(1):1–15. doi:10.3109/10409238.2013.838205

46. Nangaku M. The Heat Is On: An Expanding Role for Hypoxia-Inducible Factors in Kidney Transplantation. Journal of the American Society of Nephrology. 2007;18(1)

47. Rosenberger C, Pratschke J, Rudolph B, et al. Immunohistochemical detection of hypoxia-inducible factor-1alpha in human renal allograft biopsies. J Am Soc Nephrol. Jan 2007;18(1):343–351. doi:10.1681/asn.2006070792

48. Seino Ki, Yamauchi T, Shikata K, et al. Prevention of acute and chronic allograft rejection by a novel retinoic acid receptor-α-selective agonist. International Immunology. 2004;16(5):665–673. doi:10.1093/intimm/dxh066

49. Kiss E, Popovic ZV, Bedke J, et al. Peroxisome proliferator-activated receptor (PPAR)gamma can inhibit chronic renal allograft damage. Am J Pathol. May 2010;176(5):2150–2162. doi:10.2353/ajpath.2010.090370

50. Ashburner M, Ball CA, Blake JA, et al. Gene Ontology: tool for the unification of biology. Nature Genetics. 2000/05/01 2000;25(1):25–29. doi:10.1038/75556

51. Kanehisa M, Goto S. KEGG: Kyoto Encyclopedia of Genes and Genomes. Nucleic Acids Research. 2000;28(1):27–30. doi:10.1093/nar/28.1.27

52. Fabregat A, Jupe S, Matthews L, et al. The Reactome Pathway Knowledgebase. Nucleic Acids Research. 2018;46(D1):D649–D655. doi:10.1093/nar/gkx1132

53. Agrawal A, Balcı H, Hanspers K, et al. WikiPathways 2024: next generation pathway database. Nucleic Acids Research. 2024;52(D1):D679–D689. doi:10.1093/nar/gkad960

