## Supplemental Materials for "Total plasma cfDNA methylation in kidney transplant recipients provides insight into acute allograft rejection pathophysiology"

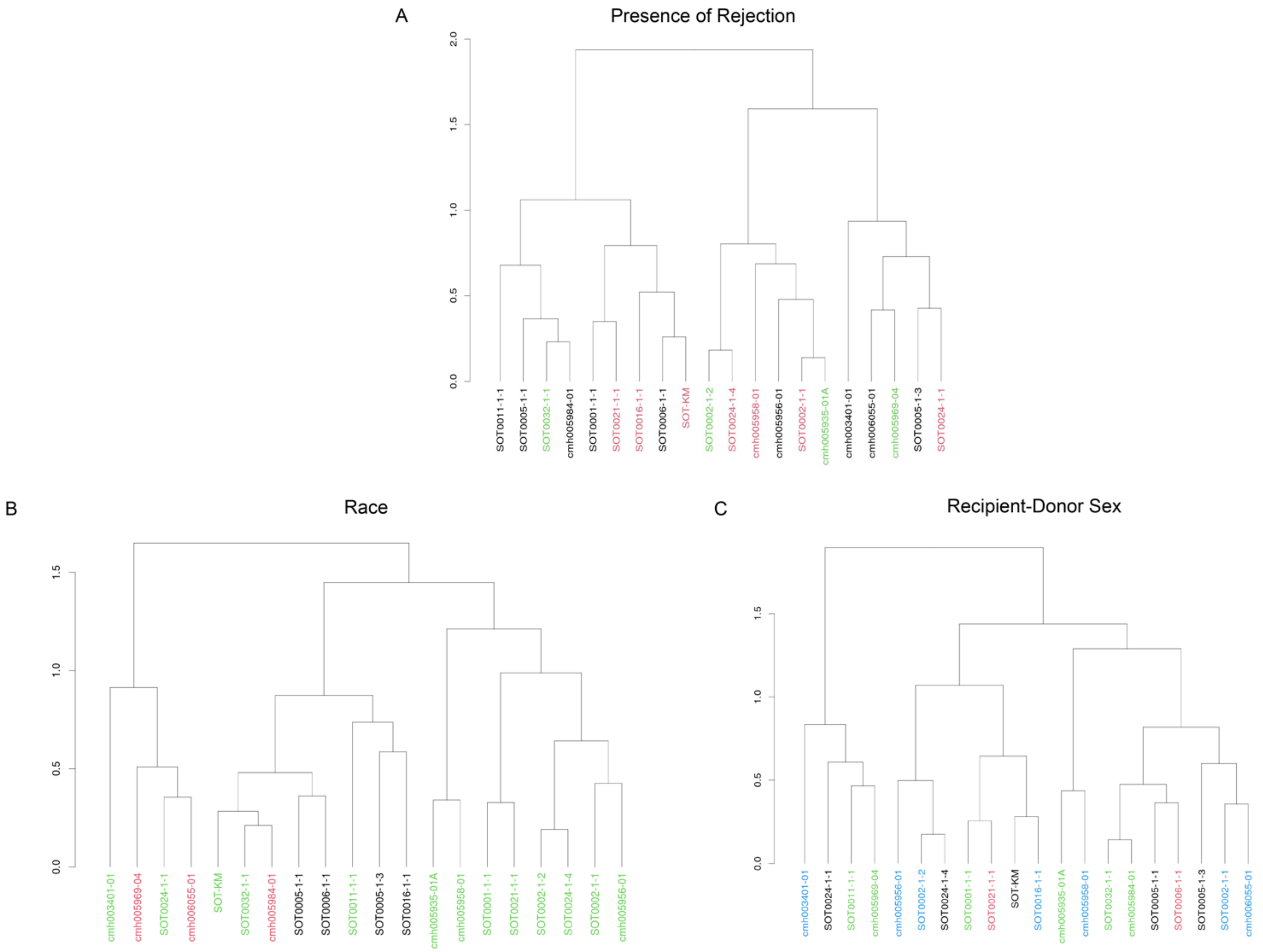


**Supplemental Figure 1 Hierarchical clustering at the top 25% most variably methylated cytosine residues shows no clustering pattern for the variables of (B) self-reported race or (C) recipient-donor sex pairing.** A) Black = Non-rejection, Red = Rejection, Green = Borderline rejection; B) Green = Caucasian, Black = Black race, Red = Non-Caucasian/Non-Black; C) in the case of recipient-donor sex pairing, Blue = male-male, Black = female-female, Green = male-female, Red = female-male.

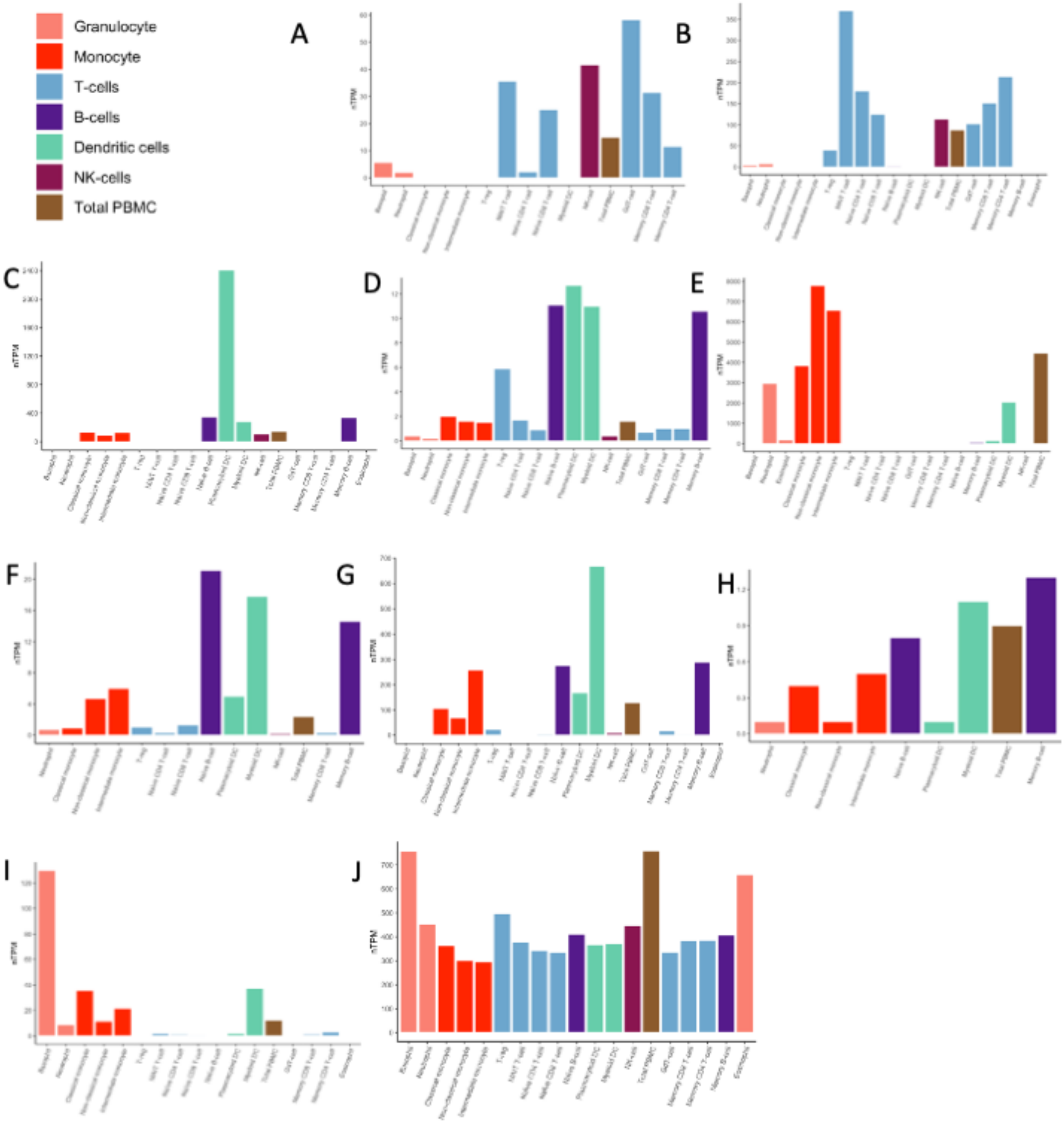


**Supplemental Figure 2 Expression levels of immune cell regulatory genes across immune cell subtypes.** A) *FASLG*, B) *IL7R*, C) *IRF8*, D) *IRF4* E) *AIF1*, F) *HLA-DOA*, G) *HLA-DQA1*, H) *HLA-DQB2*, I) *NLRP3*, J) *HMGB1*. (Normalized transcripts per million, y-axis, salmon = granulocytes, orange = monocytes, blue = T-cells, purple = B-cells, green = dendritic cells, fuchsia = natural killer (NK) cells, brown = total peripheral blood mononuclear cells (PBMC).

Image adapted from Human Protein Atlas (v23.proteinatlas.org).

**Supplemental Table 1 Percentage of DMCs per given genomic region (%) in the comparison of AAR and non-rejection. DMCs are predominately present in intergenic and intronic regions.**

| **Region** | **Percentage (%)** |
|---|---|
| Intergenic | 52.3 |
| Intron | 44.3 |
| TTS | 1.0 |
| 3' UTR | 0.7 |
| Exon | 0.7 |
| Promoter-TSS | 0.6 |
| Non-coding | 0.5 |
| 5' UTR | 0.04 |

**Supplemental Table 2 Metascape[26] annotations of hypo- and hypermethylated pathways in comparisons of AAR vs non-rejection (A, B), borderline rejection vs non-rejection (C, D), and AAR vs borderline rejection (E, F).**

**Supplemental Table 3 Enriched GO terms[50], KEGG pathways[51], Reactome gene sets[52], and WikiPathways[53] associated with immune cell regulation in the comparison of AAR vs non-rejection.** Categorization in the innate or adaptive immune response, relative methylation status (hypo- vs hypermethylation) of genes within pathway, and differentially methylated genes belonging to the pathway within our dataset are noted. Genes in bold represent genes overlapping with inflammatory response.

| Innate vs Adaptive Immunity | Methylation status | Pathway | $\log_{10}P$ | Genes |
|---|---|---|---|---|
| Innate | Hyper | innate immune response | -3.5 | ***FASLG***, ***CRP***, ***NQO1***, ***F12***, ***GCH1***, ***IRF8***, ***IFIT2***, ***PPARG***, ***CCL15***, ***SP100***, *AKAP1*, ***BCL10***, ***PJA2***, *CLEC10A*, ***USP18***, ***RPL13A***, *TRIM58*, *SAMD9*, ***NLRP2***, *ZC3HAV1*, ***ZNFX1***, ***ZDHHC11***, ***ZCCHC3***, ***IL17F***, *BPIFC*, ***RAET1G***, *BPIFB3*, ***TARM1***, ***DEFB135***, *TRIM64C* |
| | Hyper | negative regulation of mononuclear cell migration | -2.9 | ***APOD***, ***LRCH1***, ***GREM1***, ***MICOS10-NBL1*** |
| Innate, adaptive | Hyper | regulation of leukocyte activation | -3.1 | ***ABL2***, ***AHR***, ***CD80***, ***CDKN1A***, ***FANCA***, ***GLI3***, ***IL7R***, ***IDO1***, ***IRF4***, ***RORA***, ***SHH***, *SLC7A1*, ***ZEB1***, ***BCL10***, ***PJA2***, ***MERTK***, *RIPK3*, ***SHPK***, ***FBXO7***, ***GAL***, ***SAMSN1***, *KLHL25*, ***CARD11***, ***TARM1*** |
| | Hyper | regulation of leukocyte proliferation | -3 | ***AHR***, ***CD80***, ***CDKN1A***, ***CRP***, ***CSF2RB***, *HHEX*, ***IDO1***, *PTH*, ***SHH***, *SLC7A1*, *RIPK3*, ***GREM1***, ***GAL***, ***CARD11*** |
| | Hyper | mononuclear cell differentiation | -2.6 | ***CD79B***, ***CCR6***, ***FLT3***, ***GLI3***, *HHEX*, ***IRF8***, ***IL7R***, ***IRF4***, ***PPARG***, ***RORA***, ***SHH***, ***MERTK***, *RIPK3*, ***FBXO7***, ***CARD11*** |
| | Hyper | leukocyte homeostasis | -2.6 | ***F2R***, ***FLT3***, ***IL7R***, ***MERTK***, *RIPK3*, *XKR8*, *GAPT* |
| | Hyper | negative regulation of leukocyte apoptotic process | -2.3 | ***IL7R***, ***IDO1***, ***BCL10***, ***MERTK***, *PTCRA* |
| | Hyper | regulation of leukocyte migration | -2.3 | ***ABL2***, ***APOD***, ***BDKRB1***, ***CCR6***, ***EDN2***, ***SELP***, *RIPK3*, ***LRCH1***, ***GREM1***, *MTUS1*, ***MICOS10-NBL1*** |
| | Hyper | leukocyte differentiation | -2 | ***CD79B***, ***CCR6***, ***FLT3***, ***GLI3***, *HHEX*, ***IRF8***, ***IL7R***, ***IRF4***, *LBR*, ***PPARG***, ***RORA***, ***SHH***, ***MERTK***, *RIPK3*, ***FBXO7***, ***CARD11*** |
| | Hyper | regulation of leukocyte apoptotic process | -2 | ***IL7R***, ***IDO1***, ***BCL10***, ***MERTK***, *RIPK3*, *PTCRA* |
| | Hypo | MHC class II protein complex assembly | -3.3 | *HLA-DOA*, *HLA-DQA1*, *HLA-DQA2*, *HLA-DQB2* |
| | Hypo | MHC protein complex assembly | -2.9 | *HLA-DOA*, *HLA-DQA1*, *HLA-DQA2*, *HLA-DQB2* |
| | Hypo | positive regulation of leukocyte cell-cell adhesion | -2.6 | ***AIF1***, *RUNX3*, *HLA-DOA*, *HLA-DQA1*, *HLA-DQA2*, *HLA-DQB2*, ***HMGB1***, ***RPS3***, ***SHH***, *ST3GAL4*, *THY1*, ***EBI3***, ***IL36B***, ***VTCN1***, ***NLRP3*** |
| | Hypo | regulation of leukocyte chemotaxis | -2.5 | ***AIF1***, ***CSF1R***, *CYP19A1*, ***HMGB1***, *RARRES2*, ***THBS1***, *DNM1L*, *PADI2*, ***STK39*** |
| | Hypo | regulation of macrophage chemotaxis | -2.4 | ***CSF1R***, *CYP19A1*, *RARRES2*, ***THBS1*** |
| | Hypo | antigen processing and presentation of exogenous peptide antigen via MHC class II | -2.2 | *HLA-DOA*, *HLA-DQA1*, *HLA-DQA2*, *HLA-DQB2* |

| | | | | |
|---|---|---|---|---|
| | Hypo | antigen processing and presentation of peptide antigen via MHC class II | -2.1 | *HLA-DOA, HLA-DQA1, HLA-DQA2, HLA-DQB2* |
| | Hypo | positive regulation of leukocyte chemotaxis | -2.1 | ***AIF1***, ***CSF1R***, ***HMGB1***, *RARRES2*, ***THBS1***, *DNM1L*, ***STK39*** |
| | Hypo | antigen processing and presentation of peptide or polysaccharide antigen via MHC class II | -2.1 | *HLA-DOA, HLA-DQA1, HLA-DQA2, HLA-DQB2* |
| | Hypo | regulation of leukocyte migration | -2.1 | ***AIF1***, ***APOD***, ***CSF1R***, *CYP19A1*, ***HMGB1***, *RARRES2*, *ST3GAL4*, ***THBS1***, *THY1*, *DNM1L*, *PADI2*, ***STK39*** |
| | Hypo | regulation of mononuclear cell migration | -2.1 | ***AIF1***, ***APOD***, ***CSF1R***, ***HMGB1***, *RARRES2*, ***THBS1***, *PADI2*, ***STK39*** |
| | Hypo | MHC class II antigen presentation | -2 | *DYNC1I1, HLA-DOA, HLA-DQA1, HLA-DQA2, HLA-DQB2, DYNLL1, RACGAP1, TUBB1* |
| Innate, adaptive | Hypo | positive regulation of leukocyte activation | -2 | ***AIF1***, ***BCL2***, *RUNX3*, *HLA-DOA*, *HLA-DQA1*, *HLA-DQA2*, *HLA-DQB2*, ***HMGB1***, ***RPS3***, ***SHH***, ***THBS1***, *THY1*, ***EBI3***, *HAVCR1*, ***IL36B***, ***VTCN1***, ***NLRP3*** |
| | Hypo | positive regulation of macrophage chemotaxis | -2 | ***CSF1R***, *RARRES2*, ***THBS1*** |
| Adaptive | Hyper | regulation of lymphocyte activation | -3 | ***ABL2***, ***AHR***, ***CD80***, ***CDKN1A***, ***FANCA***, ***GLI3***, ***IL7R***, ***IDO1***, ***IRF4***, ***SHH***, *SLC7A1*, ***ZEB1***, ***BCL10***, ***MERTK***, *RIPK3*, ***FBXO7***, ***GAL***, ***SAMSN1***, *KLHL25*, ***CARD11***, ***TARM1*** |
| | Hyper | T cell differentiation in thymus | -2.8 | ***CCR6***, ***GLI3***, ***IL7R***, ***SHH***, *RIPK3*, ***CARD11*** |
| | Hyper | lymphocyte differentiation | -2.8 | ***CD79B***, ***CCR6***, ***FLT3***, ***GLI3***, *HHEX*, ***IRF8***, ***IL7R***, ***IRF4***, ***RORA***, ***SHH***, ***MERTK***, *RIPK3*, ***FBXO7***, ***CARD11*** |
| | Hyper | regulation of T cell migration | -2.6 | ***ABL2***, ***APOD***, ***CCR6***, *RIPK3*, ***LRCH1*** |
| | Hyper | regulation of T cell mediated cytotoxicity | -2.5 | ***IL7R***, ***TAP2***, ***STX7***, *RIPK3*, ***RAET1G*** |
| | Hyper | regulation of T cell mediated immunity | -2.5 | ***AHR***, ***CD80***, ***IL7R***, ***TAP2***, ***STX7***, *RIPK3*, ***RAET1G*** |
| | Hyper | lymph node development | -2.3 | ***IL7R***, *PDPN*, *RIPK3* |
| | Hyper | thymic T cell selection | -2.2 | ***GLI3***, ***SHH***, ***CARD11*** |
| | Hyper | lymphocyte activation | -2.2 | ***CD80***, ***CD79B***, ***CCR6***, ***FLT3***, ***GLI3***, *HHEX*, ***IRF8***, ***IL7R***, ***IDO1***, ***IRF4***, ***RORA***, ***SHH***, ***MERTK***, *RIPK3*, ***FBXO7***, ***CARD11***, *GAPT*, ***EIF2AK4*** |
| | Hyper | regulation of lymphocyte apoptotic process | -2.2 | ***IL7R***, ***IDO1***, ***BCL10***, *RIPK3*, *PTCRA* |
| | Hyper | negative regulation of lymphocyte activation | -2.2 | ***CD80***, ***GLI3***, ***IDO1***, ***SHH***, ***MERTK***, ***FBXO7***, ***GAL***, ***SAMSN1***, ***TARM1*** |
| | Hyper | negative regulation of lymphocyte apoptotic process | -2.2 | ***IL7R***, ***IDO1***, ***BCL10***, *PTCRA* |
| | Hyper | regulation of T cell apoptotic process | -2.2 | ***IL7R***, ***IDO1***, *RIPK3*, *PTCRA* |
| | Hyper | regulation of alpha-beta T cell activation | -2.1 | ***CD80***, ***GLI3***, ***IRF4***, ***SHH***, *KLHL25*, ***CARD11***, ***TARM1*** |
| | Hyper | regulation of T cell activation | -2.1 | ***ABL2***, ***CD80***, ***FANCA***, ***GLI3***, ***IL7R***, ***IDO1***, ***IRF4***, ***SHH***, *SLC7A1*, ***ZEB1***, ***BCL10***, *RIPK3*, *KLHL25*, ***CARD11***, ***TARM1*** |
| | Hyper | regulation of T cell differentiation | -2 | ***CD80***, ***FANCA***, ***GLI3***, ***IL7R***, ***IRF4***, ***SHH***, ***ZEB1***, *KLHL25*, ***CARD11*** |
| | Hyper | regulation of lymphocyte differentiation | -2 | ***CD80***, ***FANCA***, ***GLI3***, ***IL7R***, ***IRF4***, ***SHH***, ***ZEB1***, ***FBXO7***, *KLHL25*, ***CARD11*** |
| | Hypo | positive regulation of T cell activation | -2.6 | ***AIF1***, *RUNX3*, *HLA-DOA*, *HLA-DQA1*, *HLA-DQA2*, *HLA-DQB2*, ***HMGB1***, ***RPS3***, ***SHH***, *THY1*, ***EBI3***, ***IL36B***, ***VTCN1***, ***NLRP3*** |

**Supplemental Table 4 Enriched GO terms[50], KEGG pathways[51], Reactome gene sets[52], and WikiPathways[53] associated with the inflammatory response in the comparison of AAR vs non-rejection.** Relative methylation status (hypo- vs hypermethylation) of genes within pathway, and differentially methylated genes belonging to the pathway within our dataset are noted. Genes in boldface represent genes overlapping with immune cell regulation pathways.

| **Methylation Status** | Pathway | $log_{10}P$ | Genes |
|---|---|---|---|
| Hyper | response to molecule of bacterial origin | -4.5 | ***AHR***, ***FASLG***, ***BDKRB1***, ***CD80***, ***CSF2RB***, ***NQO1***, ***F2R***, *FMO1*, ***GCH1***, *HMGCS2*, ***IRF8***, *IL12RB2*, ***IDO1***, *PTGS2*, ***SELP***, ***TAP2***, ***BCL10***, ***RPL13A***, ***SHPK***, *IL37* |
| Hyper | response to bacterium | -4.2 | ***AHR***, ***FASLG***, ***BDKRB1***, ***CD80***, ***CRP***, ***CSF2RB***, ***NQO1***, ***F2R***, *FMO1*, ***GCH1***, *HMGCS2*, ***IRF8***, ***IL7R***, *IL12RB2*, ***IDO1***, *PTGS2*, ***SELP***, ***TAP2***, *THRSP*, ***BCL10***, ***USP18***, ***RPL13A***, ***SHPK***, *IL37*, ***ZNFX1***, *C15orf48*, ***IL17F***, *LYG2*, *RNASE10*, ***DEFB135*** |
| Hyper | response to lipopolysaccharide | -3.9 | ***FASLG***, ***BDKRB1***, ***CD80***, ***CSF2RB***, ***NQO1***, ***F2R***, *FMO1*, ***GCH1***, *HMGCS2*, ***IRF8***, *IL12RB2*, ***IDO1***, *PTGS2*, ***SELP***, ***BCL10***, ***RPL13A***, ***SHPK***, *IL37* |
| Hyper | positive regulation of antimicrobial peptide production | -3.6 | *KLK3*, *PGC*, ***IL17F*** |
| Hyper | positive regulation of antimicrobial humoral response | -3.4 | *KLK3*, *PGC*, ***IL17F*** |
| Hyper | regulation of antimicrobial peptide production | -3.2 | *KLK3*, *PGC*, ***IL17F*** |
| Hyper | negative regulation of immune system process | -3 | *A2M*, ***AHR***, ***APOD***, ***CD80***, ***GLI3***, ***IL7R***, ***IDO1***, ***IRF4***, *NPY5R*, ***PPARG***, ***SHH***, ***MERTK***, ***USP18***, ***LRCH1***, ***FBXO7***, ***GREM1***, ***GAL***, *UBASH3A*, ***SAMSN1***, ***TARM1***, ***MICOS10-NBL1*** |
| Hyper | Interleukin-4 and Interleukin-13 signaling | -2.9 | ***FASLG***, ***CDKN1A***, ***IRF4***, *PTGS2*, ***RORA***, ***ZEB1***, *RHOU*, ***IL17F*** |
| Hyper | Cytokine Signaling in Immune system | -2.9 | ***ABL2***, ***FASLG***, ***CD80***, ***CDKN1A***, ***CSF2RB***, ***FLT3***, *GHR*, ***IRF8***, ***IFIT2***, ***IL7R***, *IL12RB2*, ***IRF4***, *KPNA4*, *PSMB1*, *PTGS2*, ***RORA***, ***SP100***, ***ZEB1***, *OSMR*, *EIF4A3*, *NUP153*, ***USP18***, *IL37*, *IL17RB*, *NDC1*, *RHOU*, ***IL17F*** |
| Hyper | regulation of antimicrobial humoral response | -2.8 | *KLK3*, *PGC*, ***IL17F*** |
| Hyper | regulation of defense response | -2.7 | *A2M*, ***AHR***, *KLK3*, *CASP6*, ***F12***, ***FANCA***, ***IDO1***, ***IRF4***, *NPY5R*, *PGC*, ***PPARG***, *PTGS2*, ***RORA***, ***BCL10***, *OSMR*, ***PJA2***, ***USP18***, ***SHPK***, *IL37*, ***NLRP2***, *MMP26*, ***ZNFX1***, ***ZDHHC11***, ***ZCCHC3***, ***IL17F***, *PIK3AP1*, ***RAET1G***, ***EIF2AK4*** |
| Hyper | Cytokine-cytokine receptor interaction | -2.2 | ***FASLG***, ***CCR6***, ***CSF2RB***, *CSH2*, *GHR*, ***IL7R***, *IL12RB2*, ***CCL15***, *OSMR*, *IL37*, *TNFRSF19*, *IL17RB*, ***IL17F*** |
| Hyper | positive regulation of immune response | -2.2 | *KLK3*, *CASP6*, ***CD80***, ***CD79B***, *GPLD1*, ***IDO1***, *PGC*, ***TAP2***, ***STX7***, *EIF2B5*, ***BCL10***, ***PJA2***, *BTN3A2*, *BTNL2*, ***ZNFX1***, ***CARD11***, ***ZCCHC3***, ***IL17F***, *PIK3AP1*, *BTNL9*, ***RAET1G***, ***EIF2AK4*** |
| Hyper | cytokine-mediated signaling pathway | -2.1 | ***CCR6***, *ACKR2*, ***CSF2RB***, ***EDN2***, ***FLT3***, *GHR*, ***IL7R***, *IL12RB2*, ***CCL15***, ***SP100***, *OSMR*, *IL37*, *TNFRSF19*, *IL17RB*, ***IL17F*** |
| Hyper | positive regulation of receptor signaling pathway via JAK-STAT | -2.1 | *CSH2*, ***F2R***, *GHR*, ***IL7R*** |
| Hyper | regulation of cytokinesis | -2 | *INCENP*, *PIK3C3*, *OR1A2*, *BIRC6*, *FSD1*, *DCDC1* |
| Hyper | positive regulation of cytokine production | -2 | ***CD80***, ***F2R***, ***IRF8***, *IL12RB2*, ***IDO1***, ***IRF4***, *PTGS2*, ***RORA***, ***BCL10***, *TMED10*, *BTN3A2*, ***NLRP2***, ***CARD11***, ***ZCCHC3***, *ARRDC4*, ***IL17F***, *SCAMP5*, ***RAET1G*** |
| Hypo | stress-induced premature senescence | -3.2 | *PLA2R1*, *SIRT1*, *WNT16* |

| | | | |
|---|---|---|---|
| Hypo | cellular response to chemical stress | -2.9 | ***AIF1***, ***BCL2***, *GPX7, MGMT, MT3, PCNA, PPIA*, ***RPS3***, *NET1, PLA2R1, SIRT1, STAU2*, ***STK39***, *WNT16*, ***NLRP3***, *MPV17L* |
| Hypo | cytokine-mediated signaling pathway | -2.4 | *TNFRSF17*, ***CSF1R***, *ACSL1, XCR1, IL5RA, LIFR, LIMS1, MT3, TRAF3, IL18R1*, ***EBI3***, *CCL26, SIRT1*, ***IL36B***, *IL37*, ***STK39***, *CARD14, ADIPOR2* |
| Hypo | Inflammatory response pathway | -2.3 | *FN1, IL5RA, LAMC1*, ***THBS1*** |
| Hypo | positive regulation of cytokine production | -2.1 | ***AIF1***, ***CSF1R***, *FLT4, GDF2*, ***HMGB1***, *LY9, MMP12, P2RX7*, ***RPS3***, ***THBS1***, *TRAF3, IL18R1*, ***EBI3***, *PLA2R1, SIRT1, CADM1, CHIA, SULF2*, ***VTCN1***, ***NLRP3***, *SCIMP* |

**Supplemental Table 5 Enriched GO terms[50], KEGG pathways[51], Reactome gene sets[52], and WikiPathways[53] associated with lipid metabolism in the comparison of AAR vs non-rejection.** Relative methylation status (hypo- vs hypermethylation) of genes within pathway, and differentially methylated genes belonging to the pathway within our dataset are noted.

| **Methylation Status** | Pathway | $\log_{10}p$ | Genes |
|---|---|---|---|
| Hyper | Fatty Acids and Lipoproteins Transport in Hepatocytes | -4.6 | *ACKR2, DBI, DPYSL2, CTTN, GHR, HMGCS1, HMGCS2, LBR, LOXL2, PIK3C3, SHH, HIP1R, RABEP1, CLEC10A, AAK1, RHOU, RAB17, DENND1C, ACAD11, NOSTRIN, SCARA5* |
| Hyper | Metabolism of Lipid metabolisms | -4.5 | *AHR, ALOX12, DBI, FDXR, FHL2, HMGCS1, HMGCS2, LBR, NRF1, PIK3C3, PPARG, PTGS2, RORA, SC5D, SCD, THRSP, CYP4F2, PLA2G4C, SPTLC2, TECR, LPGAT1, MED6, MORC2, NCOA6, GPD1L, TIAM2, LPIN3, PIP4K2C, ACAD11, ACBD5, PTGR2, NUDT7* |
| Hyper | PPARA activates gene expression | -4 | *AHR, FHL2, HMGCS1, HMGCS2, NRF1, PPARG, RORA, MED6, NCOA6, TIAM2* |
| Hyper | Regulation of Lipid metabolism metabolism by PPARalpha | -3.9 | *AHR, FHL2, HMGCS1, HMGCS2, NRF1, PPARG, RORA, MED6, NCOA6, TIAM2* |
| Hyper | Fatty acid metabolism | -3.7 | *ALOX12, DBI, PTGS2, SCD, THRSP, CYP4F2, TECR, MORC2, ACAD11, ACBD5, PTGR2, NUDT7* |
| Hyper | Cholesterol metabolism with Bloch and Kandutsch-Russell pathways | -2.7 | *HMGCS1, HMGCS2, LBR, SC5D, SCD* |
| Hyper | Lipid metabolism biosynthetic process | -2.4 | *ALDH3B2, ALOX12, EDN2, FDXR, HMGCS1, HMGCS2, LBR, PIK3C3, PTGS2, SC5D, SCD, ST8SIA2, PLA2G4C, TM9SF2, SPTLC2, TECR, LPGAT1, ABCA8, SCCPDH, LPIN3, PIP4K2C, PNPLA1* |
| Hyper | unsaturated fatty acid metabolic process | -2.4 | *ALOX12, EDN2, PTGS2, SCD, CYP4F2, PLA2G4C, PTGR2* |
| Hyper | fatty acid metabolic process | -2.3 | *ALOX12, DBI, EDN2, PTGS2, SCD, CYP4F2, PLA2G4C, TECR, MORC2, LPIN3, ACAD11, ACBD5, PTGR2, NUDT7* |
| Hyper | Cholesterol synthesis disorders | -2.3 | *HMGCS1, LBR, SC5D* |
| Hyper | cholesterol biosynthetic process | -2.2 | *HMGCS1, HMGCS2, LBR, SC5D* |
| Hyper | Lipid metabolism metabolism in senescent cells | -2.2 | *CDKN1A, PTGS2, PLA2G4C* |
| Hyper | unsaturated fatty acid biosynthetic process | -2.2 | *ALOX12, EDN2, PTGS2, SCD* |
| Hyper | positive regulation of Lipid metabolism transport | -2.1 | *FASLG, DBI, PPARG, CYP4F2, ABCA8, GAL* |
| Hypo | Lipid metabolism transport | -5 | *APOD, APOH, CES1, CETP, CFTR, CIDEA, ACSL1, GOT2, P2RX7, PLTP, ABCD4, RBP1, APOL1, ABCA10, ATP11B, ANO7, PCTP, PLA2G2F, APOL5, APOL3, SLC22A9, ABCA13, SLC22A24* |
| Hypo | Cholesterol metabolism | -4.6 | *APOH, CETP, PLTP, VDAC1, VAPB, ANGPTL4, NCEH1, PCSK9* |
| Hypo | positive regulation of Lipid metabolism transport | -4.4 | *CES1, CETP, CYP19A1, ACSL1, P2RX7, PLTP, PLA2R1, SIRT1, C1QTNF1, ABCA13* |
| Hypo | Lipid metabolism localization | -4.3 | *APOD, APOH, CES1, CETP, CFTR, CIDEA, ACSL1, GOT2, P2RX7, PLTP, ABCD4, RBP1, APOL1, ABCA10, ATP11B, ANO7, PCTP, PLA2G2F, APOL5, APOL3, SLC22A9, ABCA13, SLC22A24* |
| Hypo | positive regulation of Lipid metabolism localization | -4.1 | *CES1, CETP, CIDEA, CYP19A1, ACSL1, P2RX7, PLTP, PLA2R1, SIRT1, C1QTNF1, ABCA13* |
| Hypo | regulation of Lipid metabolism transport | -4.1 | *CES1, CETP, CYP19A1, ACSL1, P2RX7, PLTP, SHH, THBS1, PLA2R1, SIRT1, C1QTNF1, ABCA13, PCSK9* |
| Hypo | regulation of Lipid metabolism localization | -3.9 | *CES1, CETP, CIDEA, CYP19A1, ACSL1, P2RX7, PLTP, SHH, THBS1, PLA2R1, SIRT1, C1QTNF1, ABCA13, PCSK9* |
| Hypo | Fatty Acids and Lipoproteins Transport in Hepatocytes | -3.9 | *ACAT2, APOH, CES1, CETP, CFTR, CSNK1G3, ACSL1, HMGCS2, LGALS3BP, PLTP, ABCD4, SHH, VDAC1, APOL1, DNM1L, PLA2R1, SH3BP4, CSNK1G1, TMPRSS4, RHOJ, NOSTRIN, PCSK9* |
| Hypo | Metabolism of Lipid metabolisms | -2.7 | *ACAT2, CGA, CIDEA, CYP2C19, CYP2D6, CYP2E1, CYP19A1, AKR1C2, ACSL1, HMGCS2, LHB, CH25H, VAPB, PLA2R1, DHRS7B, ANGPTL4,* |

| | | | |
|---|---|---|---|
| | | | *ETNK2, SPTLC3, PI4K2A, AHRR, PCTP, PLA2G2F, PIP4K2C, MED28, SELENOI, CYP2U1, PLA2G4E, NEU4, ACOT12, ACSM6, CERS3* |
| Hypo | positive regulation of cholesterol transport | -2.7 | *CES1, CETP, PLTP, SIRT1, ABCA13* |
| Hypo | Lipid metabolism biosynthetic process | -2.6 | *CES1, CFTR, CYP2D6, CYP2E1, CYP19A1, ACSL1, HMGCS2, LHB, P2RX7, PIGH, RBP1, ST3GAL4, CH25H, EFR3A, DHRS7B, A4GALT, ETNK2, SPTLC3, PI4K2A, PIP4K2C, ST6GALNAC5, SELENOI, GGTLC1, IP6K3, ACSM6, CERS3* |
| Hypo | regulation of cholesterol transport | -2.5 | *CES1, CETP, PLTP, SHH, SIRT1, ABCA13, PCSK9* |
| Hypo | fatty acid transport | -2.2 | *ACSL1, GOT2, P2RX7, ABCD4, RBP1, PLA2G2F, SLC22A9* |
| Hypo | cholesterol metabolic process | -2.2 | *CES1, CETP, CFTR, CYP2D6, HMGCS2, APOL1, CH25H, PCSK9* |
| Hypo | Cholesterol metabolism | -2.1 | *ACAT2, APOH, CETP, PLTP, VDAC1, PCSK9* |
| Hypo | regulation of fatty acid transport | -2.1 | *ACSL1, P2RX7, THBS1, PLA2R1* |
| Hypo | Familial hyperlipidemia type 1 | -2.1 | *CETP, PLTP, ANGPTL4* |
| Hypo | regulation of cholesterol efflux | -2 | *CES1, CETP, PLTP, SHH, SIRT1* |

**Supplemental Table 6 Enriched GO terms[50], KEGG pathways[51], Reactome gene sets[52], and WikiPathways[53] associated with the kynurenine-tryptophan pathway in the comparison of AAR vs non-rejection.** Relative methylation status (hypo- vs hypermethylation) of genes within pathway, and differentially methylated genes belonging to the pathway within our dataset are noted.

| **Methylation Status** | **Pathway** | **$\log_{10}p$** | **Genes** |
|---|---|---|---|
| Hyper | Kynurenine pathway and links to cell senescence | -4.1 | *AHR*, *CDKN1A*, *IDO1*, *ACMSD*, *EIF2AK4* |
| Hyper | Tryptophan metabolism (WikiPathway) | -3.4 | *AHR*, *DDC*, *IDO1*, *TPH1*, *ACMSD* |
| Hyper | Tryptophan metabolism (KEGG Pathway) | -2 | *DDC*, *IDO1*, *TPH1*, *ACMSD* |

**Supplemental Table 7A Enriched transcription factor binding motifs among relatively hypomethylated DMCs in plasma cfDNA in AAR vs non-rejection**

| Motif | Name | p-value | q-value | Target sequences with motif (%) | Background sequences with motif (%) |
|---|---|---|---|---|---|
| 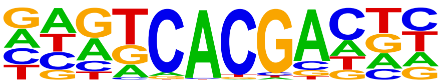 | Npas4 | 1x10$^{-10}$ | <0.0001 | 9.63 | 8.28 |
| 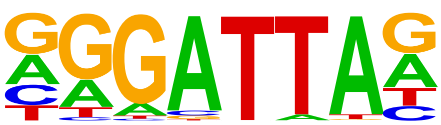 | GSC | 1x10$^{-8}$ | <0.0001 | 24.41 | 22.65 |
| 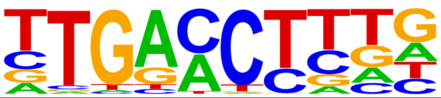 | RARα | 1x10$^{-8}$ | <0.0001 | 33.57 | 31.65 |
| 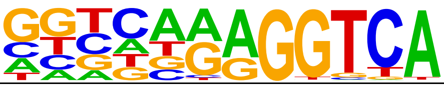 | COUP-TFII | 1x10$^{-6}$ | <0.0001 | 14.70 | 13.44 |
| 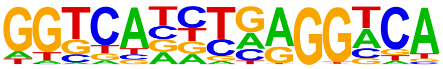 | THRα | 1x10$^{-6}$ | <0.0001 | 5.48 | 4.69 |
| 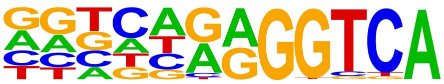 | EAR2 | 1x10$^{-5}$ | 0.0001 | 13.55 | 12.43 |
| 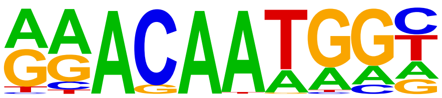 | Sox15 | 1x10$^{-5}$ | 0.0001 | 13.26 | 12.15 |
| 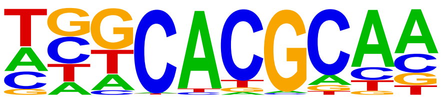 | Arnt:Ahr | 1x10$^{-5}$ | 0.0001 | 5.33 | 4.61 |
| 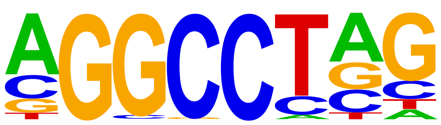 | ZNF711 | 1x10$^{-5}$ | 0.0001 | 12.02 | 10.97 |
| 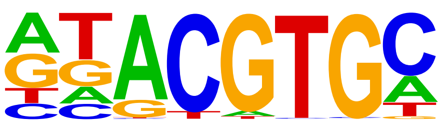 | HIF-1β | 1x10$^{-5}$ | 0.0003 | 10.85 | 9.90 |

**Supplemental Table 7B Enriched transcription factor binding motifs among relatively hypermethylated DMCs in plasma cfDNA in AAR vs non-rejection**

| Motif | Name | p-value | q-value | Target sequences with motif (%) | Background sequences with motif (%) |
|---|---|---|---|---|---|
| 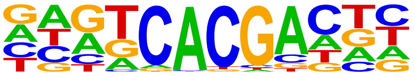 | Npas4 | $1x10^{-11}$ | <0.0001 | 10.06 | 8.49 |
| 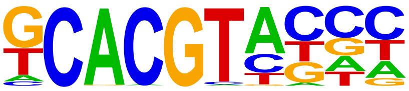 | HIF2α | $1x10^{-9}$ | <0.0001 | 3.56 | 2.72 |
| 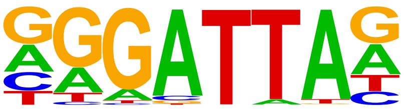 | GSC | $1x10^{-8}$ | <0.0001 | 25.34 | 23.28 |
| 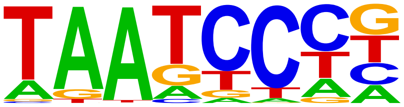 | Pitx1 | $1x10^{-8}$ | <0.0001 | 57.68 | 55.39 |
| 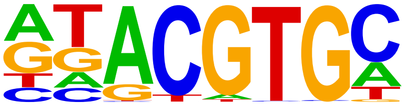 | HIF-1β | $1x10^{-7}$ | <0.0001 | 10.98 | 9.61 |
| 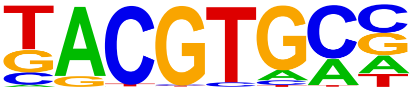 | HIF-1α | $1x10^{-7}$ | <0.0001 | 2.66 | 2.00 |
| 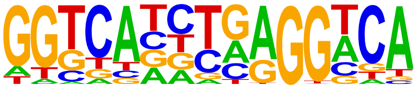 | THRα | $1x10^{-7}$ | <0.0001 | 5.42 | 4.47 |
| 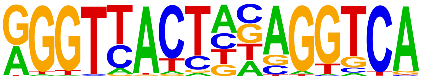 | LXRE | $1x10^{-7}$ | <0.0001 | 2.60 | 1.97 |
| 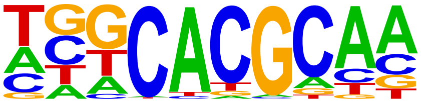 | Arnt:Ahr | $1x10^{-5}$ | 0.0001 | 5.21 | 4.41 |
| 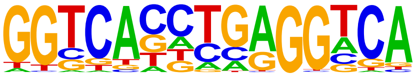 | THRβ | $1x10^{-5}$ | 0.0001 | 5.70 | 4.90 |
